# Diagnostic Performance of MRI Radiomics for Classification of Alzheimer’s disease, Mild Cognitive Impairment, and Normal Subjects: A Systematic Review and Meta-analysis

**DOI:** 10.1101/2023.03.26.23287754

**Authors:** Ramin Shahidi, Mansoureh Baradaran, Ali Asgarzadeh, Sara Bagherieh, Zohreh Tajabadi, Akram Farhadi, Setayesh Sotoudehnia Korani, Mohammad Khalafi, Parnian Shobeiri, Hamidreza Sadeghsalehi, Arezoo Shafieyoun, Mohammad Amin Yazdanifar, Aparna Singhal, Houman Sotoudeh

## Abstract

**Background:** Alzheimer’s disease (AD) is a debilitating neurodegenerative disease. Early diagnosis of AD and its precursor, mild cognitive impairment (MCI), is crucial for timely intervention and management. Magnetic resonance imaging (MRI) radiomics showed a promising result for diagnosing and classifying AD, and MCI from normal subjects. Thus, we aimed to systematically evaluate the diagnostic performance of the MRI radiomics for this task.

**Methods and materials:** A comprehensive search of the current literature was conducted using relevant keywords in PubMed/MEDLINE, Embase, Scopus, and Web of Science databases from inception to October 17, 2022. Original studies discussing the diagnostic performance of MRI Radiomics for the classification of AD, MCI, and normal subjects were included. Method quality was evaluated with the Quality Assessment of Diagnostic Accuracy Studies (QUADAS-2), and the Radiomic Quality Score tool (RQS).

**Results:** We identified 10 studies that met the inclusion criteria, involving a total of 3446 participants. The overall quality of the included studies was moderate to high. The pooled sensitivity and specificity of MRI radiomics for differentiating AD from normal subjects were 0.8822 (95% CI 0.7888-0.9376), and 0.8849 (95% CI 0.7978-0.9374), respectively. The pooled sensitivity and specificity of MRI radiomics for differentiating MCI from normal subjects were 0.7882 (95% CI 0.6272-0.8917) and 0.7736 (95% CI 0.6480-0.8639), respectively. Also, the pooled sensitivity and specificity of MRI radiomics for differentiating AD from MCI were 0.6938 (95% CI 0.6465-0.7374) and 0.8173 (95% CI 0.6117-0.9270), respectively.

**Conclusion:** MRI radiomics has promising diagnostic performance in differentiating AD, MCI, and normal subjects. It can potentially serve as a non-invasive and reliable tool for early diagnosis and classification of AD and MCI.

## 1. Introduction

Alzheimer’s disease (AD) is the most common progressive neurodegenerative disorder with a high socioeconomic burden and morbidity (Lane, Hardy, & Schott, 2018). Clinically, AD is defined primarily by comprehensive dementia, which includes memory disorder, cognitive disorder, executive dysfunction, personality, and behavior abnormalities, and is accompanied by mental disorder symptoms in the majority of patients (Lyketsos et al., 2011). AD and other dementias grew in incidence and prevalence by 147.95 and 160.84%, respectively, from 1990 to 2019 (Li et al., 2022). AD and other dementias are expected to affect 152 million individuals by 2050 (Nichols et al., 2022). Although meticulous care and medicine might temporarily alleviate these symptoms, no definite strategies exist to prevent or cure AD (Srivastava, Ahmad, & Khare, 2021). Mild cognitive impairment (MCI), a stage between normal aging and dementia, has been recognized as a significant risk factor for AD. According to epidemiological studies, roughly 10-12% of people with MCI develop AD each year (Langa & Levine, 2014). Since many elderly individuals suffer from MCI but do not fulfill the diagnostic criteria for AD, early care for people at this stage may successfully prevent disease progression (C. R. Jack et al., 2013).

MCI has little effect on everyday activities, and those affected have normal cognitive performance (Vega & Newhouse, 2014). However, MCI is characterized by variable cognitive performance and clinical progression, and the clinical results are unknown. Some MCI patients stay stable or even return to normal function, while others develop AD (Gauthier et al., 2006). Therefore, there is an urgent need to find biomarkers that may identify and predict high-risk MCI patients who will proceed to AD, since these patients will need intervention. Cerebrospinal fluid (CSF) chemical alterations and neuroimaging evaluations of brain structure and function have been established as viable biomarkers of AD (Da et al., 2014; Dickerson, Wolk, & Initiative, 2013; Salvatore, Cerasa, & Castiglioni, 2018). These characteristics include a rise in CSF tau, hypometabolism in the posterior cingulate, and atrophy of the hippocampi (Geuze, Vermetten, & Bremner, 2005; Sperling et al., 2011). The automated diagnosis and prognosis of AD patients using a machine-learning model and the combined use of these biomarkers have been shown to be reliable and highly accurate (Salvatore, Battista, & Castiglioni, 2016). Due to the high incidence of AD, the expensive expense of these procedures, and their relative complexity of usage, the application of these biomarkers may be restricted.

Radiomics is a novel technique based on the in-depth fusion of computer science and medicine. It represents the variability of disorder via imaging properties and is both inexpensive and non-invasive (Lambin et al., 2017; Yip & Aerts, 2016). During the early years, radiomics was extensively used in oncology (Gillies, Kinahan, & Hricak, 2016), and it is now used for the diagnosis and categorical assessment of MCI and AD (Feng & Ding, 2020). In recent years, neuroimaging studies have shown that white matter (WM) degradation and demyelination in the microscopic and macroscopic structure of WM are crucial physiological markers for identifying AD risk and disease development (Nasrabady, Rizvi, Goldman, & Brickman, 2018). These microstructural alterations may be observed in three-dimensional whole-brain WM radiomics investigations (Shao et al., 2018). In addition, grey matter (GM) atrophy and pathological alterations in the cerebrospinal fluid (CSF) may detect very early disturbances associated with pathological aging and AD (C. R. Jack, Jr. et al., 2018).

To date, the current systematic review and meta-analysis is the first to present the status of published literature on the diagnostic performance of MRI radiomics for diagnosing AD and MCI. Also, the potential values of MRI radiomics in predicting AD progression and development of AD in individuals with MCI are discussed.

## 2. Methods and materials

This systematic review was conducted in accordance with the Preferred Reporting Items for Systematic Reviews and Meta-analysis (PRISMA) statement (Page et al., 2021). The protocol of study has been registered with the international prospective register of systematic reviews (PROSPERO) under the code CRD42023389087.

### 2.1. Search strategy

A comprehensive literature review was performed to identify observational studies discussing the diagnostic performance of MRI Radiomics in Alzheimer’s disease (AD) and mild cognitive impairment (MCI) diagnosis. Four online databases, including PubMed/MEDLINE, Embase, Scopus, and Web of Science were systematically searched from inception to October 17, 2022, using relevant keywords. These terms included “Alzheimer disease”, “Alzheimer”, “Alzeimer”, “Primary Senile Degenerative Dementia”, “Senile Dementia”, “Presenile Dementia”, “Mild Cognitive Impairment”, “Radiomic”, and “Radiomics”. A search strategy was designed for each database using a combination of these keywords with appropriate Boolean operators (OR/AND). The search strategy used in each database is summarized in more detail in Appendix 1. No restrictions in terms of publication time, study design, language, and country of publication were applied to retrieve all available literature.

### 2.2. Inclusion and exclusion criteria

All of the observational studies, including cross-sectional, case-control, and cohort studies that addressed the diagnostic performance of MRI radiomics in AD and MCI diagnosis, which met the inclusion criteria were enrolled in this study. The inclusion criteria were as follows: (1) patient’s age ≥ 18 years; (2) AD and MCI diagnosis confirmed by mini-mental state examination (MMSE) score; (3) containing original data regarding brain structural and/or functional abnormalities on brain MRI; and (4) using radiomics methods for AD and MCI diagnosis with reported diagnostic performance indices, such as sensitivity and specificity.

The following studies were excluded: (1) other types of studies rather than observational (including case reports/series, editorials, comments, correspondence, guideline, experimental, and interventional studies, as well as meta-analysis, and systematic and narrative reviews); (2) grey literature; (3) articles without available full text; (4) irrelevant studies; (5) studies that did not report diagnostic performance indices, such as sensitivity and specificity; (6) studies that focused on the prediction of the MCI progression to AD; (7) studies that merged MCI and AD patient to differentiate them from cognitively normal controls; and (8) pre-prints and not peer-reviewed publications.

### 2.3. Study selection process

Extracted citations were imported into the EndNote 20 software (Clarivate Analytics, Philadelphia, PA, USA), and duplicates were removed. Two independent reviewers (A.A and MA.Y) carried out the first level of screening by selecting eligible studies based on their titles and abstracts. Selected articles were subjected by the same reviewers to the second level of screening to be reviewed by their full text based on the inclusion and exclusion criteria. Any disagreement was dissolved by consulting a third reviewer (R.S).

### 2.4. Data extraction

The whole manuscripts of the final included studies were reviewed by two independent investigators (M.K and S.S), and the following information was collected using a predefined Microsoft Excel worksheet: first author’s name, period of each study, the country where the study was conducted, dataset source, total number, age, gender, and MMSE score of healthy controls and patients with AD and MCI, features of the used MRI method (strength, vendor, and sequence), type of segmentation, ROI (segmented location) / VOI, radiomics feature extraction and selection (number of features, extracted features, software, feature extraction, and selection method), pre-processing status, and statistical data of radiomics findings (TP, TN, FP, and FN). In cases where TP, TN, FP, and FN are not reported, we will reconstruct these four items based on the sensitivity and specificity reported as well as the number of controls and patients. We will round these four items closer to whole numbers if they are in decimal form. Consequently, there may be a slight difference between the sensitivity and specificity mentioned in our article and those in the included articles due to this rounding.

### 2.5. Quality assessment

Three researchers (M.K, S.S and A.S) independently assessed the methodological quality of the included studies with the Quality Assessment of Diagnostic Accuracy Studies (QUADAS-2) (Schueler, Schuetz, & Dewey, 2012), and the Radiomic Quality Score tool (RQS) (Zhong et al., 2021). QUADAS_J2 tool was developed to evaluate the quality of non-randomized studies and contains four domains: patient selection, index test, reference standard, and flow and timing. Each domain is appraised in terms of the risk of bias. Patient selection, index tests, and reference standards are also appraised in terms of concerns regarding the applicability and classify the risk of bias of each included a study into low, high, or unclear risk. Signaling questions are included to help judge the risk of bias. The RQS that analyzes the quality of a radiomics study is a consensus list composed of sixteen items for methodological issues specific to radiomics studies for a maximum score of 36.

### 2.6 Quantitative Meta-Analysis

#### 2.6.1 Software

R version 4.2.2 (R Core Team [2022]. R: A language and environment for statistical computing. R Foundation for Statistical Computing, Vienna, Austria) was used for all calculations, visualizations, and further analysis on meta-analyses with substantial heterogeneity.

#### 2.6.2 Statistical analysis

The statistical significance level was defined as a p-value of < 0.05. The effect size was quantified using the proportion of true and false cases that were correctly identified as such by the AI model. The analytical model was composed of fixed and random effects, simultaneously. The I^2^ index was used to determine heterogeneity, with an I^2^ < 40% value indicating that inconsistency across studies is not significant. In such cases, we conducted the meta-analysis using the fixed effects model. However, in case the I^2^ estimates varied by more than 40%, we performed the analysis using the random effects technique.

#### 2.6.3 Sensitivity analysis

To further illuminate the possible sources of heterogeneity, we performed a sensitivity analysis on meta-analyses with considerable heterogeneity (I^2^ > 40%). We removed one study each time and recalculated the effect size (Leave-One-Out Analyses).

## 3. RESULTS

### 3.1. Overview & Basic Information

234 records were identified through database searching. After removing the duplicates, 147 records remained, out of which 87 were screened. Eventually, the full-text versions of 67 articles were obtained and carefully evaluated, leading to the inclusion of 10 articles in the final analysis (Figure 1).

**Figure 1.**
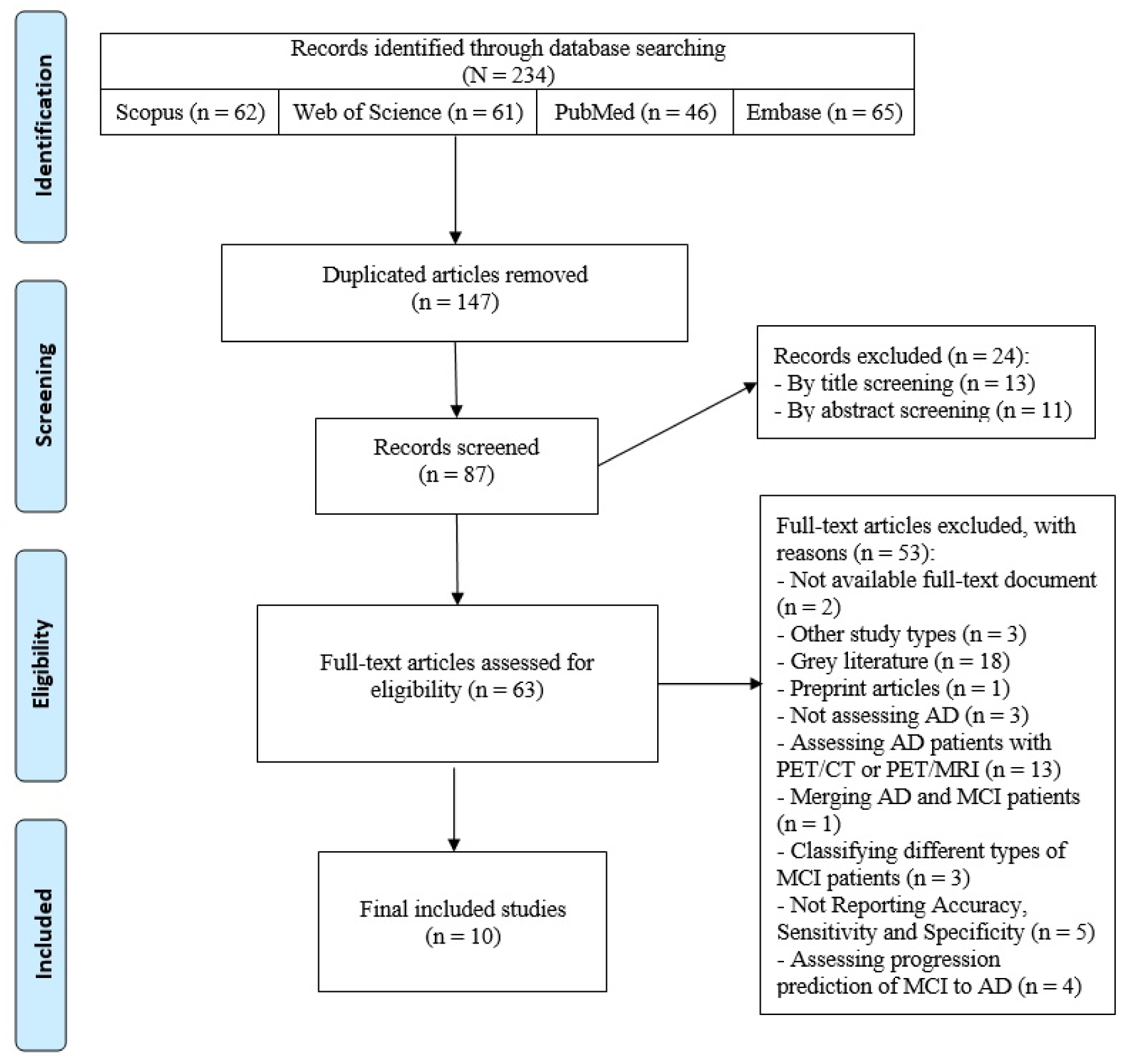
Flow diagram summarizing the selection of eligible studies.

Table 1 summarizes the demographic and neuropsychological data of 979, 1300, and 1123 individuals with AD, MCI, and CN, respectively, adding up to a total of 3446 Individuals, with mean ages of 71.27 (SD: 9.15), 71.8 (SD: 8.92), and 68.2 (SD: 7.74), respectively.

**Table 1.** Demographic and neuropsychological data of the included individuals.

MMSE: Mini-Mental State Examination
|  | AD |  |  |  | MCI |  |  |  | CN |  |  |  |
| --- | --- | --- | --- | --- | --- | --- | --- | --- | --- | --- | --- | --- |
| First Author | Number | Age | Gender (M/F) | MMSE | Number | Age | Gender (M/F) | MMSE | Number | Age | Gender (M/F) | MMSE |
| Cheung, 2022 | 97 | NA | NA | NA | 293 | NA | NA | NA | 192 | NA | NA | NA |
| Du, 2022 | 36 | 72.45±2.8 | 17/19 | 22.5±3.0 | NA | NA | NA | NA | 36 | 72.08±1.4 | 17/19 | 29.2±0.4 |
| F. Feng, 2018 | 38 | 71.7±8.3 | 16/22 | 17.6±5.6 | 33 | 70.6±8.2 | 14/19 | 26.6±2.6 | 45 | 68.2±6.9 | 22/23 | 28.6±1.4 |
| Q. Feng, 2018 | 78 | 69.18±12.23 | 25/53 | 16.94±5.94 | NA | NA | NA | NA | 44 | 65.43±9.7 | 20/24 | 29.14±0.77 |
| Q. Feng, 2019 | NA | NA | NA | NA | 42 | 67.14±10.57 | 18/24 | 25.88±0.92 | 44 | 65.43±9.7 | 20/24 | 29.14±0.77 |
| Q. Feng, 2020 | 97 | 67.67±10.73 | 39/58 | 17.42±5.73 | 53 | 65.13±13.48 | 26/27 | 26.00±0.96 | 45 | 64.76±9.92 | 20/25 | 29.11±0.78 |
| liu, 2022 | 80 | 65(46-88) | 42/38 | 15(0-25) | NA | NA | NA | NA | 80 | 64.5(48-83) | 40/40 | 28(12-30) |
| Ranjbar, 2019 | 41 | 76.1±8.7 | 16/25 | NA | 70 | 76.0±8.4 | 43/27 | NA | 62 | 75.2±4.7 | 26/36 | NA |
| Zhao, 2020 | 261 | 68.84±8.17 | 107/154 | 16.53±5.22 | 223 | 68.91±8.94 | 105/118 | 24.94±3.22 | 231 | 66.93±6.91 | 97/134 | 28.46±1.64 |
| Zheng, 2022 | 251 | 74.82±7.49 | 131/120 | 23.2±2.11 | 636 | 73.28±7.69 | 376/260 | 27.44±1.82 | 388 | 74.62±5.73 | 198/190 | 29.09±1.07 |

Table 2, on the other hand, summarizes the basic characteristics of the included studies. Accordingly, 5 studies utilized the Alzheimer’s Disease Neuroimaging Initiative database (ADNI), while 5 other used their centers’ private databases. Furthermore, two studies, both of which had ADNI as a source of data, used a second database, as well. The segmentation process was performed automatically, manually, and both automatically/manually in 6, 2, and 2 studies, respectively. Also, being the selected region of interest (ROI) in 6 out of the 10 included articles, hippocampus was the most frequently named ROI of choice among the articles.

**Table 2.**
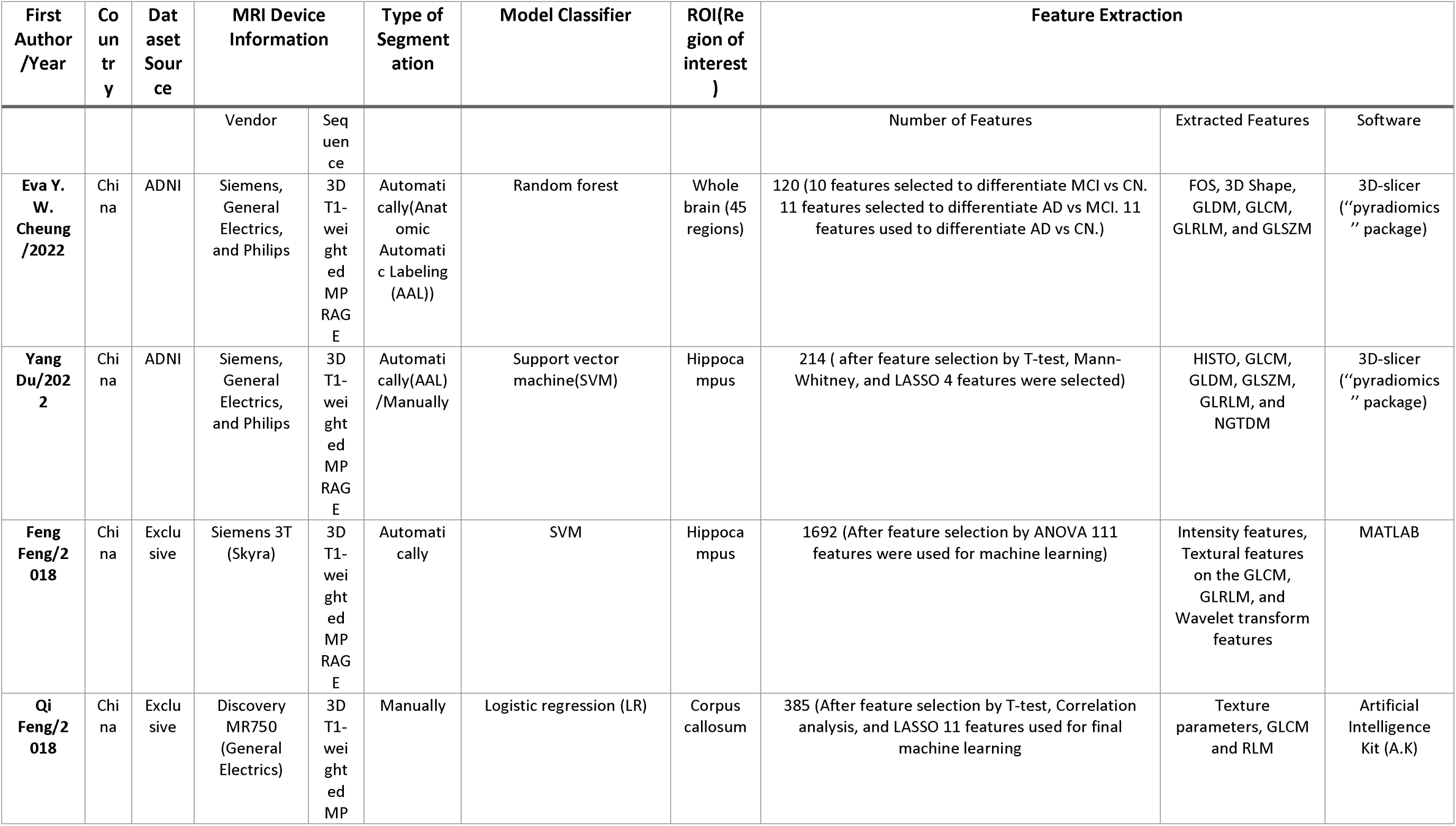
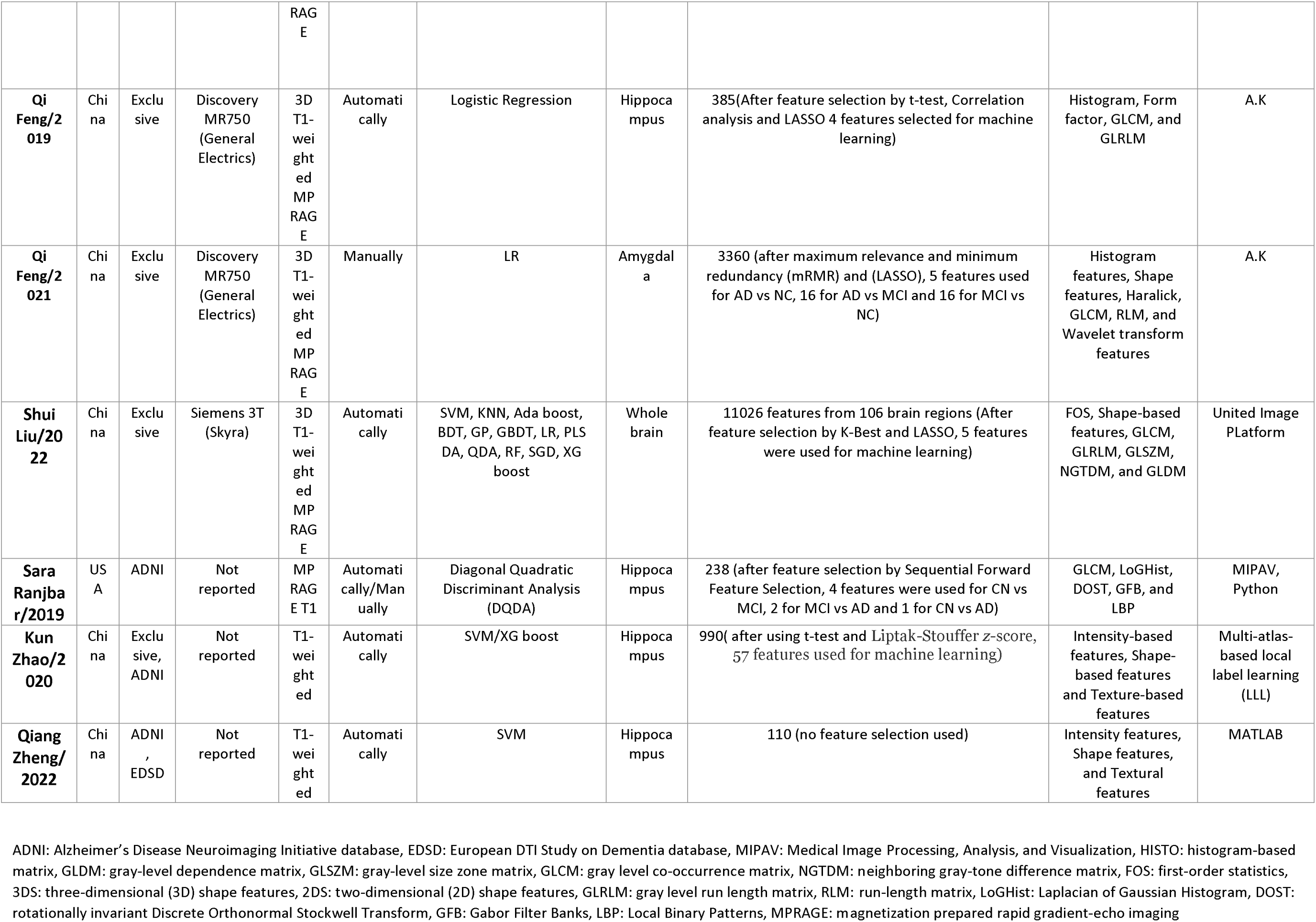
Basic characteristics of the included studies.

The results of the critical appraisal step based on two separate tools, i.e., the QUADAS and RQS tools, revealed low concerns regarding the applicability aspects of the obtained data (Supplementary Figure 1, Supplementary Figure 2, and Supplementary Figure 3). In the risk of bias domain, all the studies achieved relatively decent scores in all the fields, and “Flow and Timing” turned out to be the strong suit of all the included studies. However, study conducted by Cheung et al. (Cheung, Chau, Tang, & Life, 2022) was a major source of concern regarding reference standards (Supplementary Figure 1, Supplementary Table 1, and Supplementary Table 2).

The pooled sensitivity, specificity, and precision of the proposed AI models were meta-analyzed. Moreover, the mean and standard deviation (SD) of the accuracy and AUCs of the included articles are reported below.

### 3.2. AD Vs. MCI

6 studies provided data on the diagnostic accuracy metrics of radiomics models in differentiating AD from MCI cases. The pooled sensitivity, specificity, precision, accuracy, and AUC of the mentioned task were 0.6938 (95% CI: [0.6465; 0.7374], I^2 = 44.0%, I^2 95% CI: [0.0%; 77.8%]) (Figure 2), 0.8173 (95% CI: [0.6117; 0.9270], I^2 = 91.3%, I^2 95% CI: [83.7%; 95.3%]) (‘Figure 3), 0.7468 (95% CI: [3.06; 5.35], I^2 = 93.9%, I^2 95% CI: [89.3%; 96.5%]) (Figure 4), 74.22 (SD: 13.63), and 77.52 (SD: 11.39), respectively. Since the I^2^ level was considerably high in specificity and precision analyses, a sensitivity analysis was conducted, indicating the lowest heterogeneity is achieved by removing Cheung et al. study (Cheung, et al., 2022) (Figure 5), and Zheng et al. study (Zheng, Zhang, Li, Tong, & Ouyang, 2022) (Figure 6), respectively. Despite an I^2 level of 44%, none of the studies was detected as an outlier for sensitivity meta-analysis.

**Figure 2.**
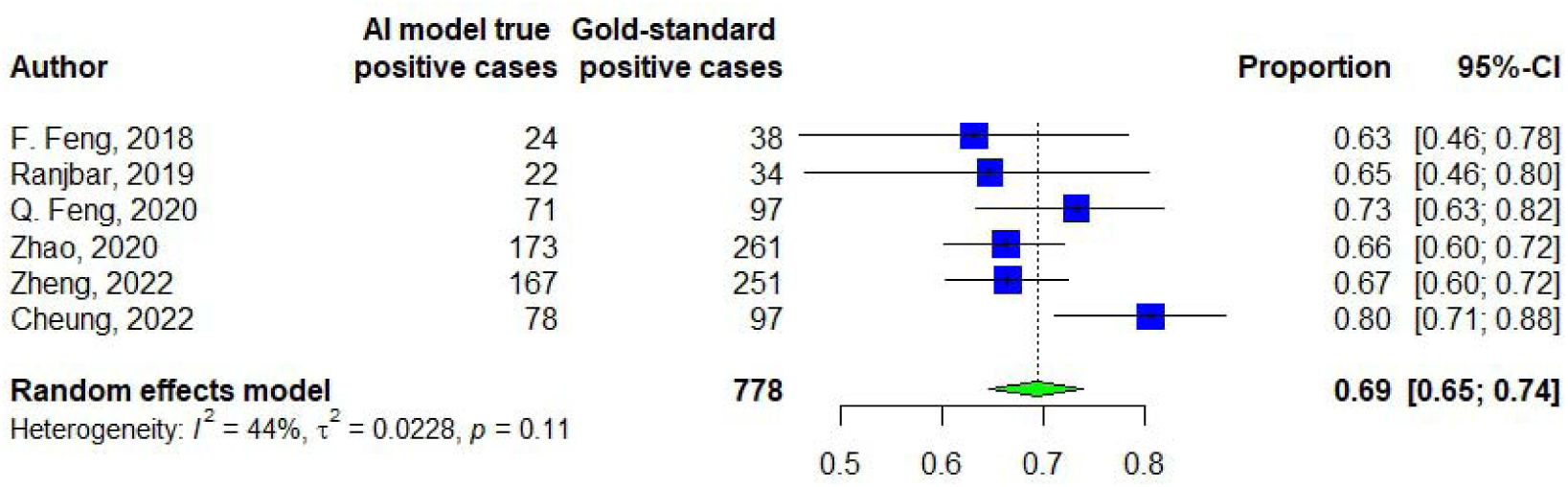
Sensitivity of differentiating AD from MCI.

**Figure 3.**
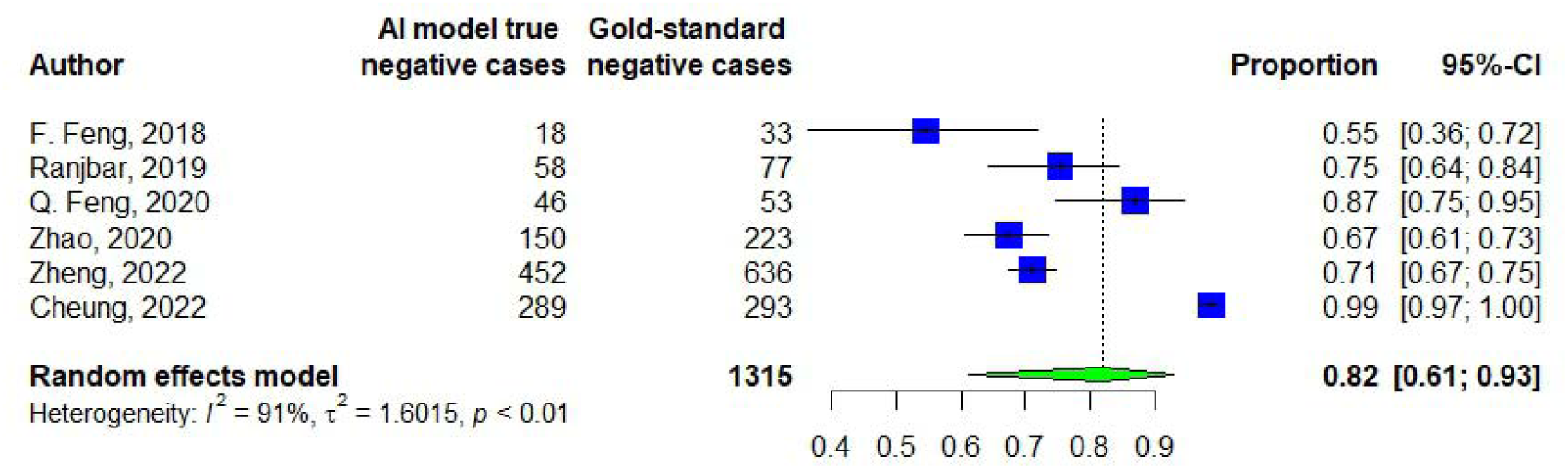
Specificity of differentiating AD from MCI.

**Figure 4.**
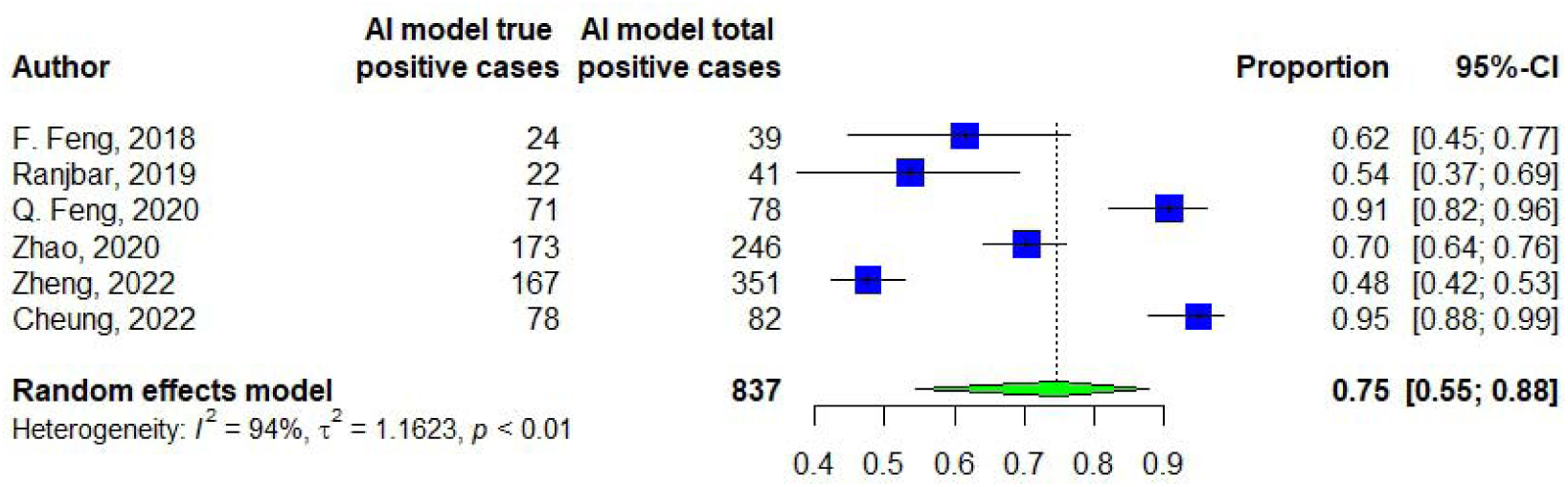
Precision of differentiating AD from MCI.

**Figure 5.**
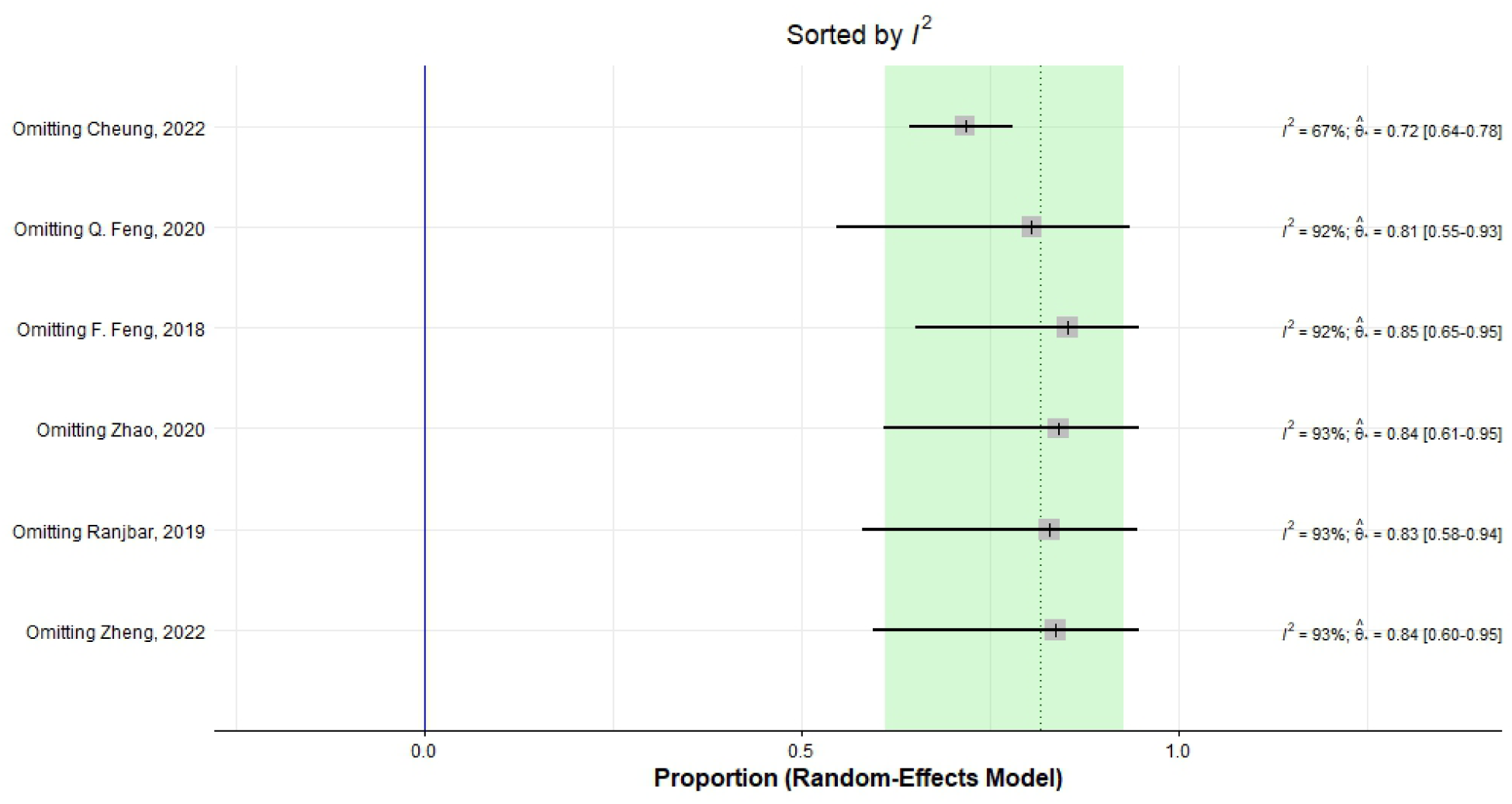
Heterogeneity of specificity of differentiating AD from MCI.

**Figure 6.**
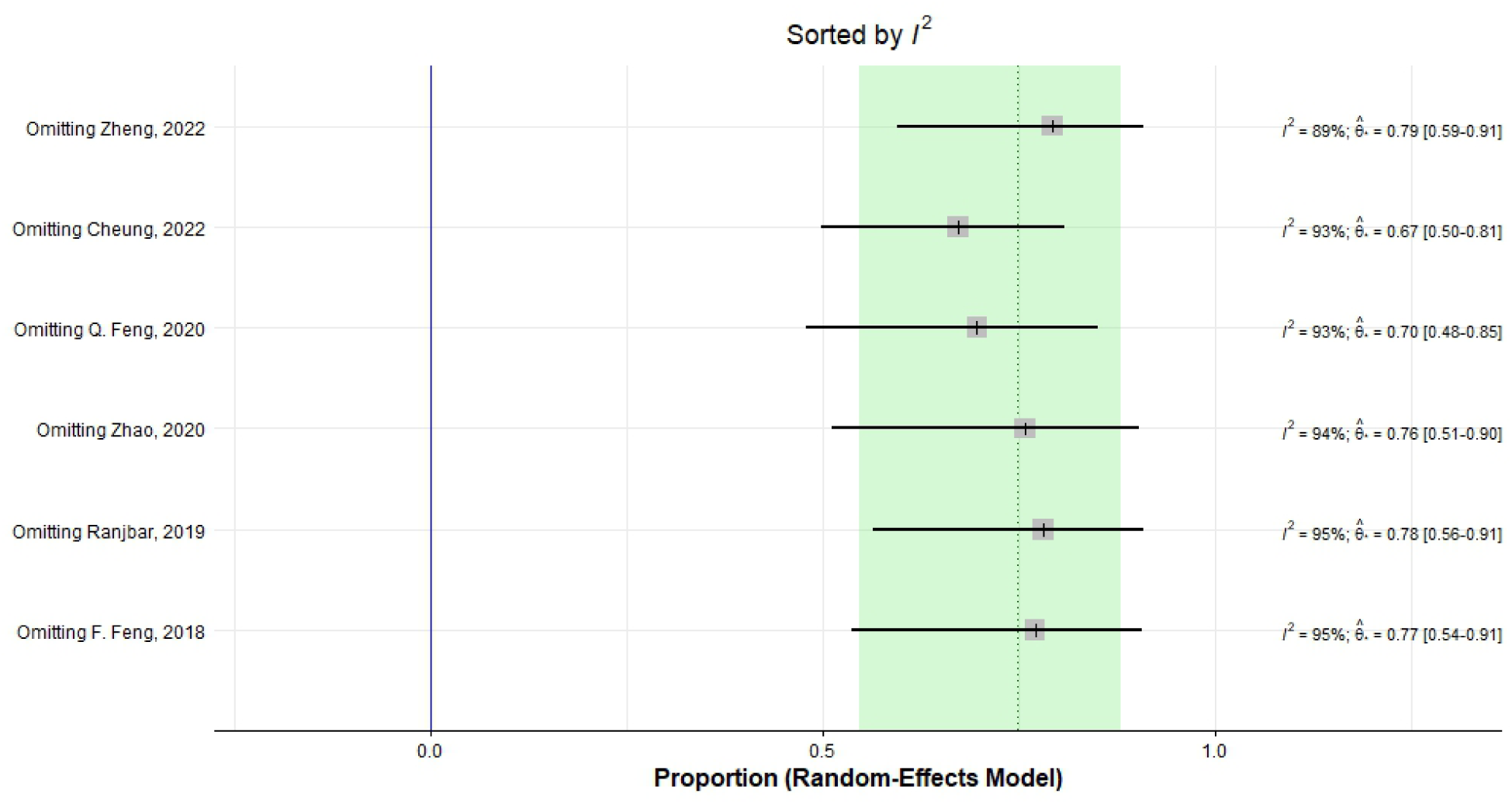
Heterogeneity of precision of differentiating AD from MCI.

### 3.3. AD Vs. CN

8 studies provided data on the diagnostic accuracy metrics of radiomics models in differentiating AD from CN cases. The pooled sensitivity, specificity, precision, accuracy, and AUC of the mentioned task were 0.8822 (95% CI: [0.7888; 0.9376], I^2 = 72.6%, I^2 95% CI: [44.0%; 86.6%]) (Figure 7), 0.8849 (95% CI: [0.7978; 0.9374], I^2 = 89.2%, I^2 95% CI: [81.1%; 93.8%]) (Figure 8), 0.8779 (95% CI: [0.8255; 0.9161], I^2 = 68.2%, ], I^2 95% CI: [33.2%; 84.8%]) (Figure 9), 87.70 (SD: 0.08), and 90.42 (SD: 0.08) respectively. Since the I^2^ level was considerably high in sensitivity, specificity, and precision analyses, a sensitivity analysis was conducted, indicating the lowest heterogeneity is achieved by removing Zheng et al. study (Zheng, et al., 2022) (Figure 10), Q. Feng et al. study (Q. Feng et al., 2018) (Figure 11), and Q. Feng et al. study (Q. Feng, et al., 2018) (Figure 12), respectively.

**Figure 7.**
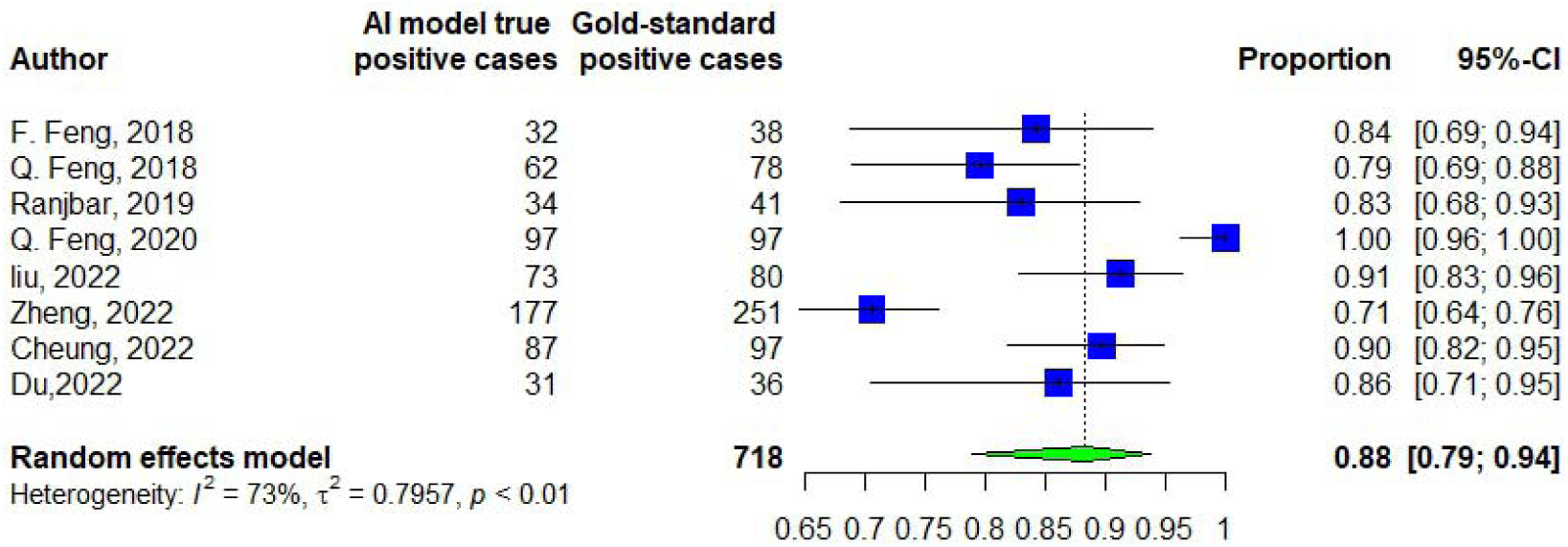
Sensitivity of differentiating AD from CN.

**Figure 8.**
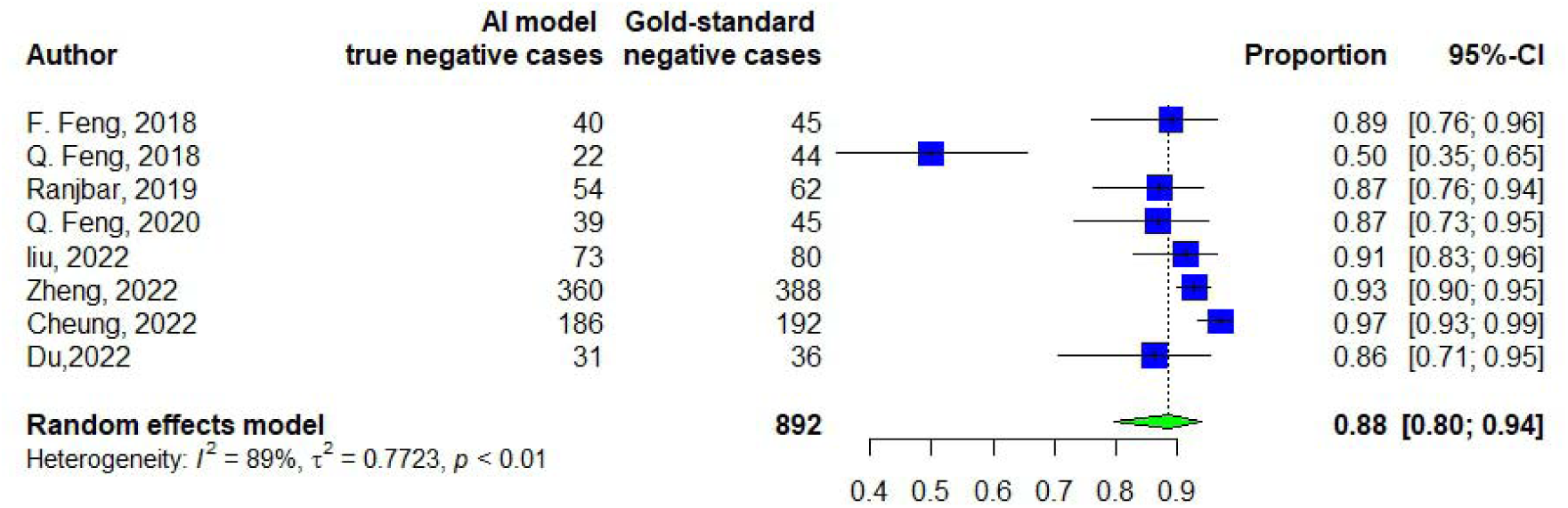
Specificity of differentiating AD from CN.

**Figure 9.**
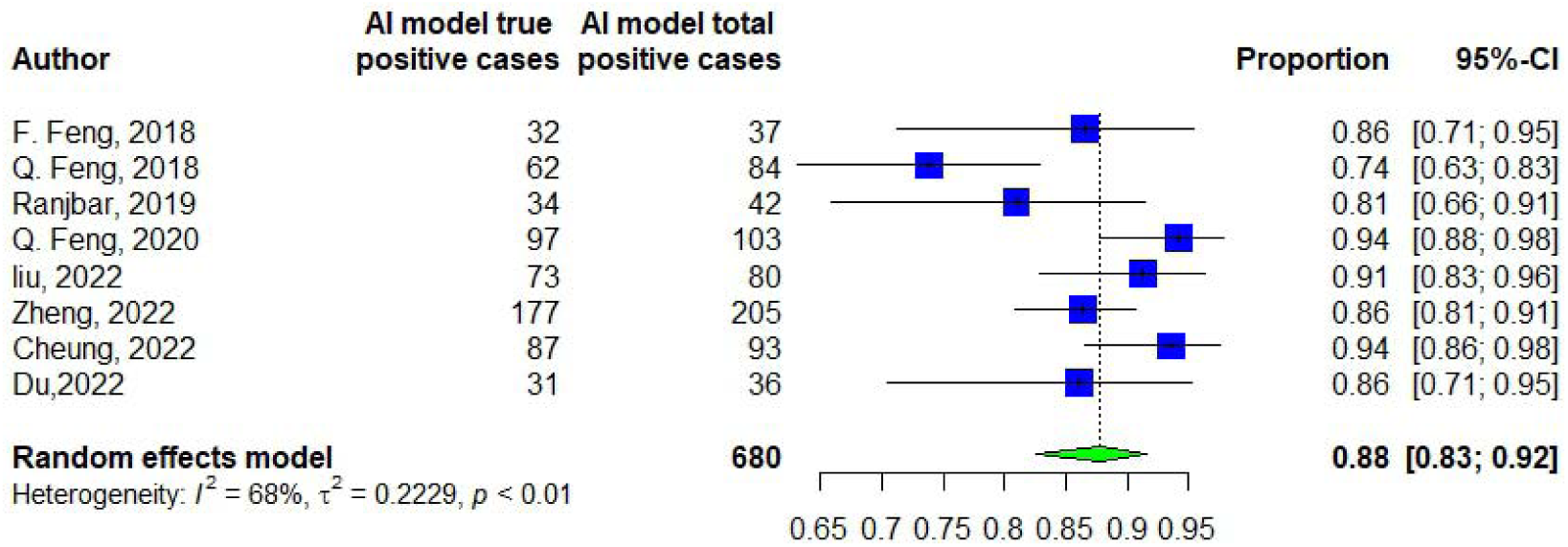
Precision of differentiating AD from CN.

**Figure 10.**
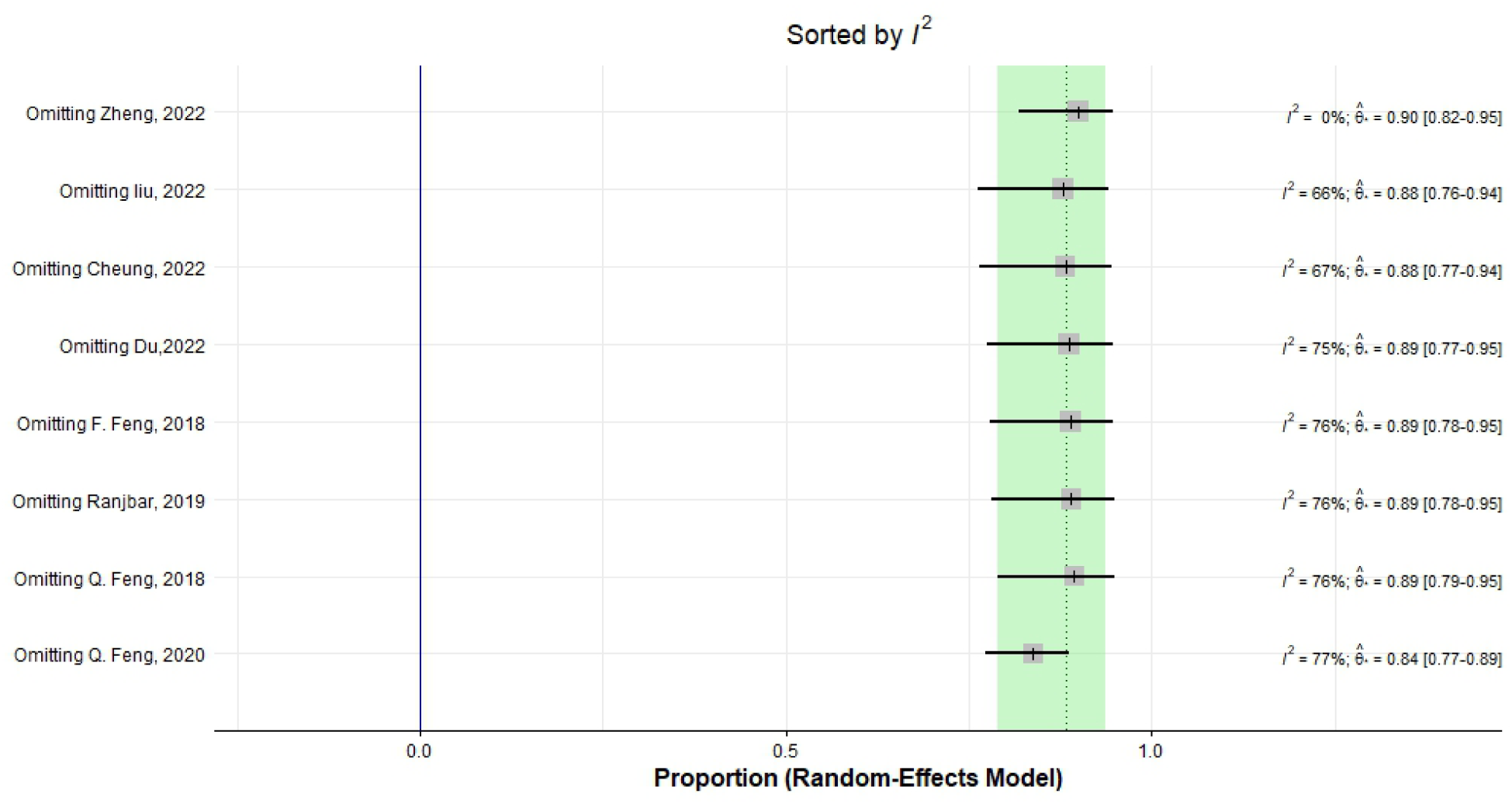
Heterogeneity of sensitivity of differentiating AD from CN.

**Figure 11.**
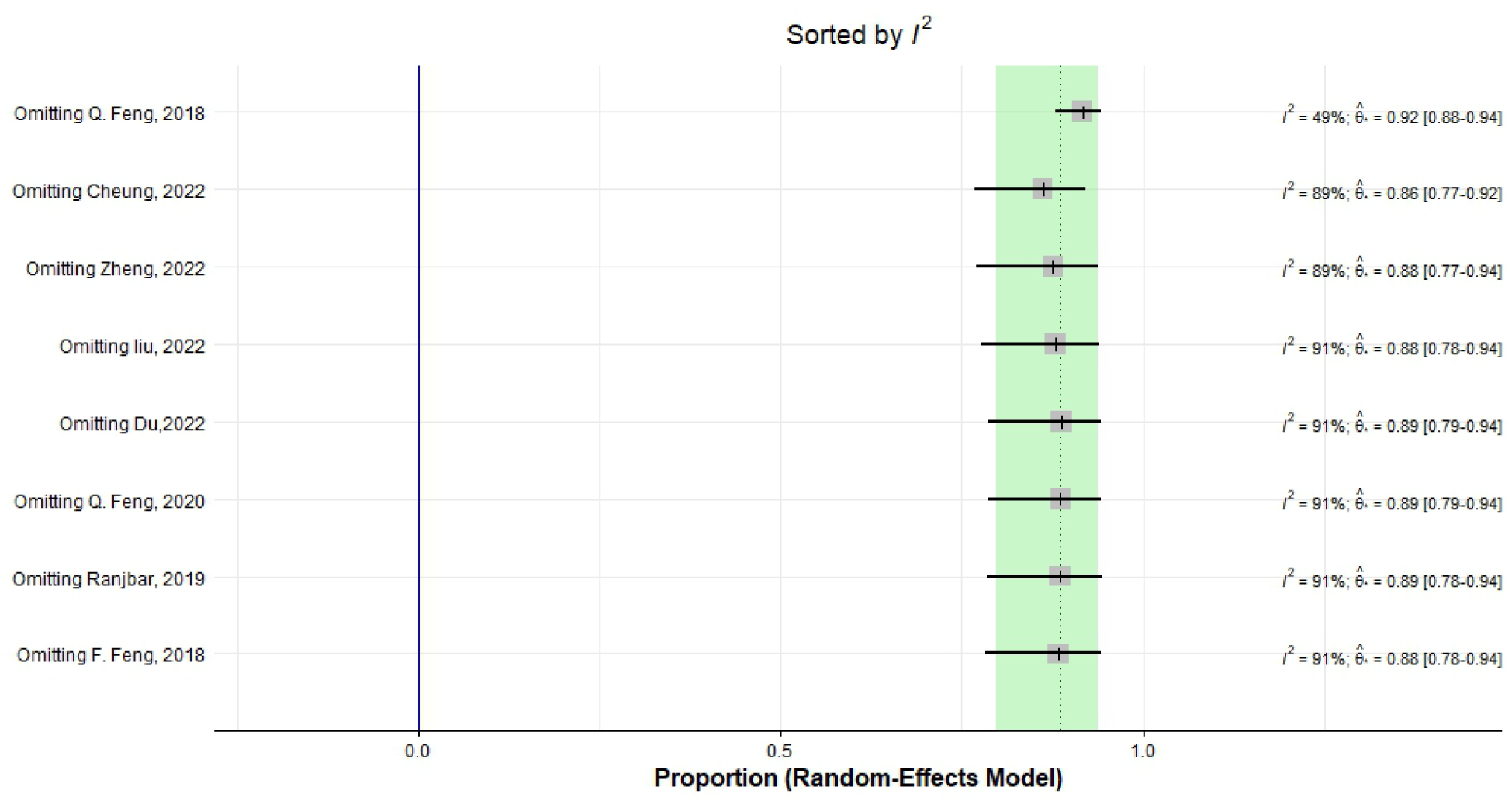
Heterogeneity of specificity of differentiating AD from CN.

**Figure 12.**
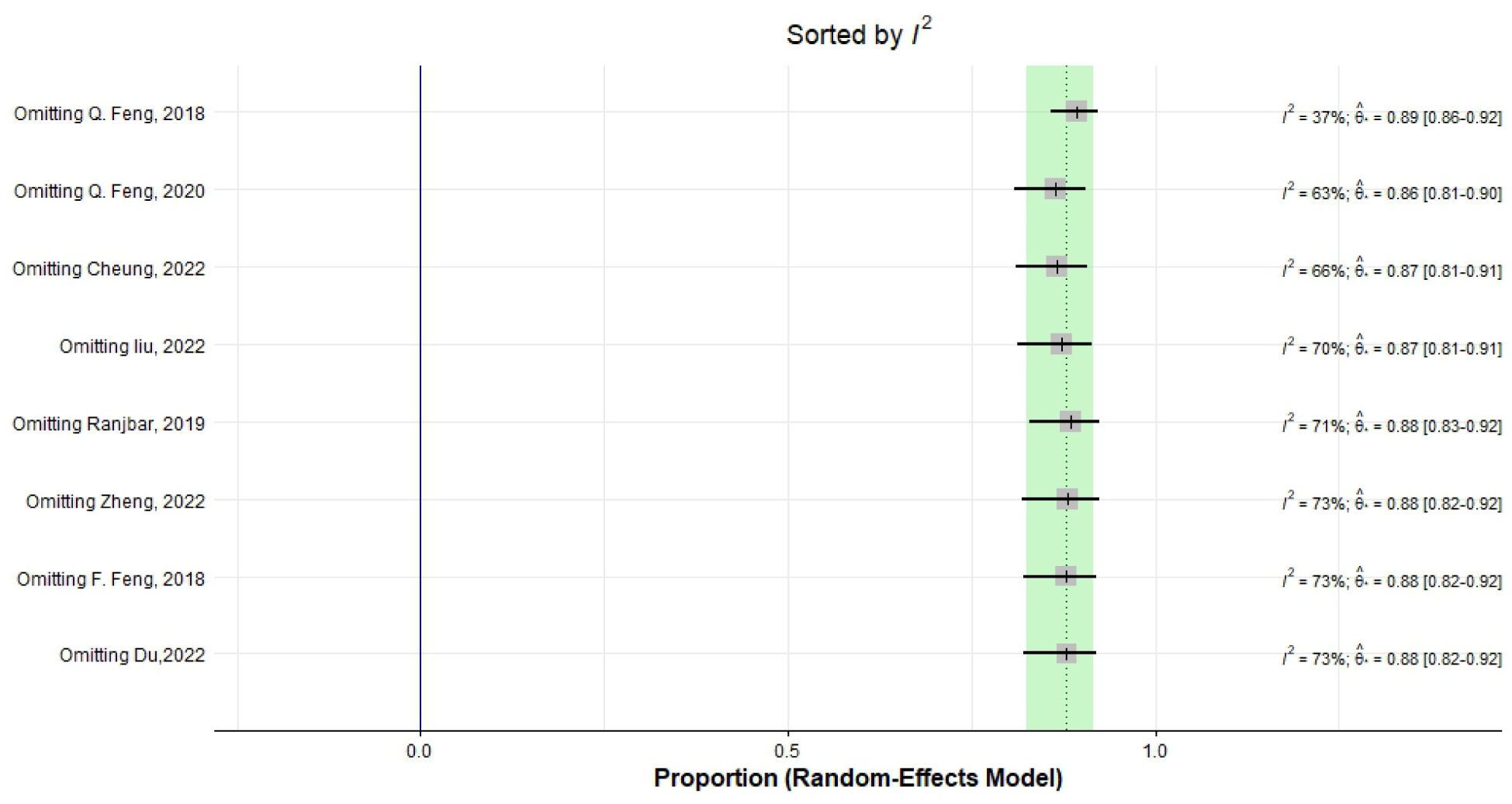
Heterogeneity of precision of differentiating AD from CN.

### 3.4. MCI Vs. CN

6 studies provided data on the diagnostic accuracy metrics of radiomics models in differentiating MCI from CN cases. The pooled sensitivity, specificity, precision, accuracy, and AUC of the mentioned task were 0.7882 (95% CI: [0.6272; 0.8917], I^2 = 91.2%, I^2 95% CI: [83.7%;95.3%]) (Figure 13), 0.7736 (95% CI: [0.6480; 0.8639], I^2 = 94.3%, I^2 95% CI: [90.1%;96.7%]) (Figure 14), 0.797 (95% CI: [0.6793; 0.8791], I^2 = 93.9%, I^2 95 % CI: [89.3%; 96.5%]) (Figure 15), 76.6 (SD: 11.65), and 79.91 (SD: 11.09), respectively. Since the I^2^ level was considerably high in sensitivity, specificity, and precision analyses, a sensitivity analysis was conducted, indicating the lowest heterogeneity is achieved by removing Cheung et al. study (Cheung, et al., 2022) (Figure 16), Zhao et al. study (Zhao et al., 2020) (Figure 17), and Cheung et al. study (Cheung, et al., 2022) (Figure 18), respectively.

**Figure 13.**
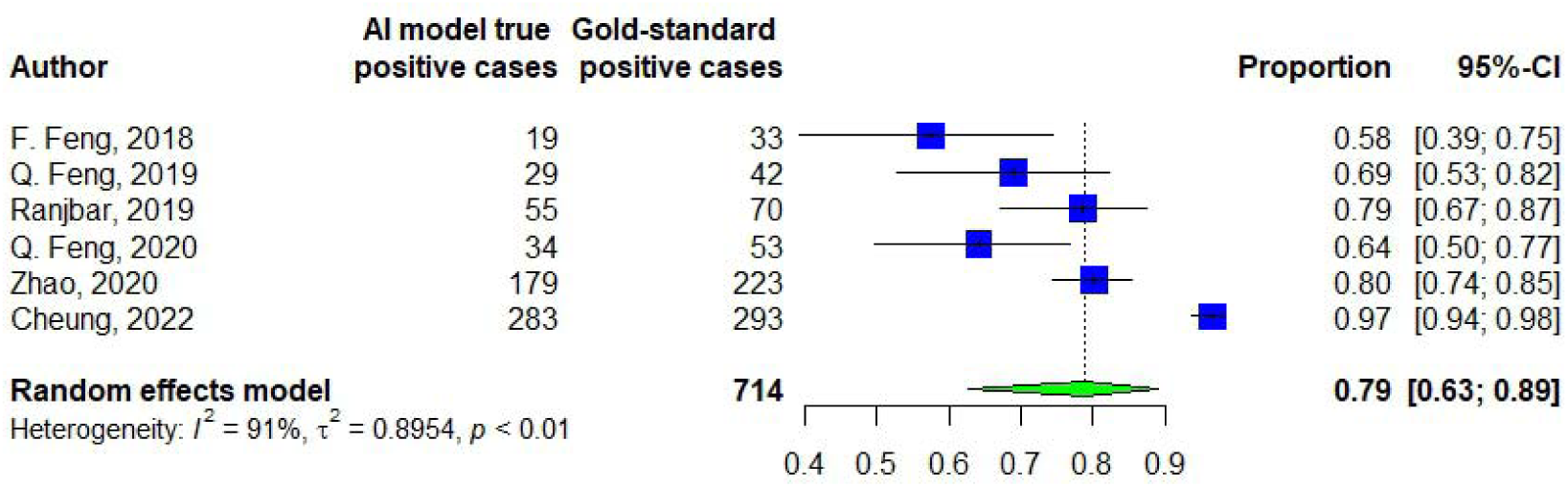
Sensitivity of differentiating CN from MCI.

**Figure 14.**
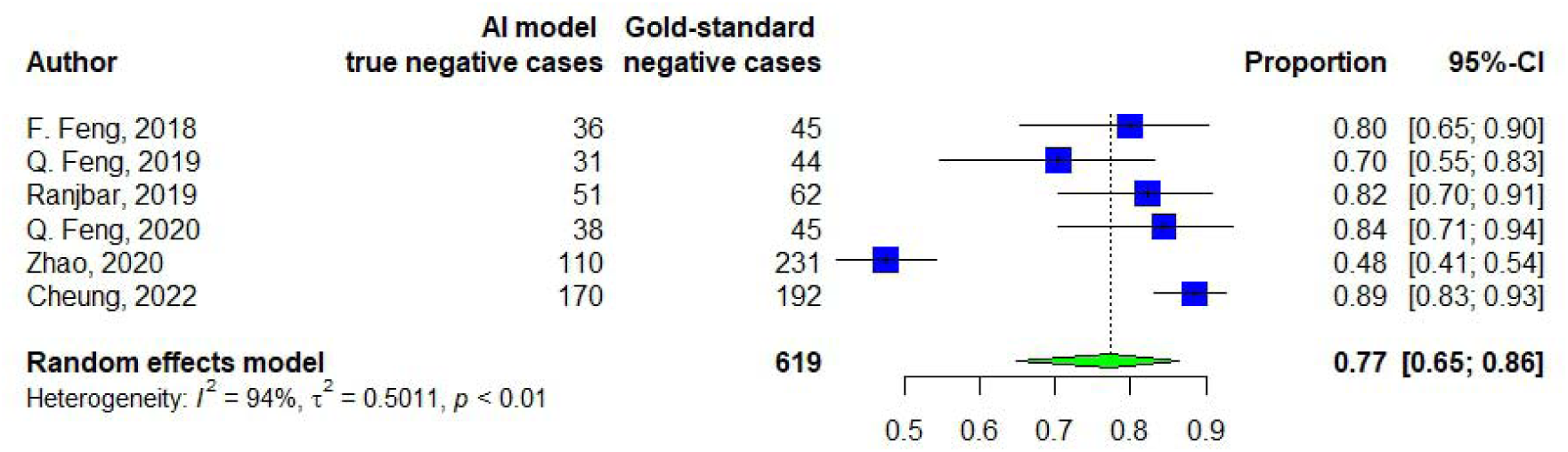
Specificity of differentiating CN from MCI.

**Figure 15.**
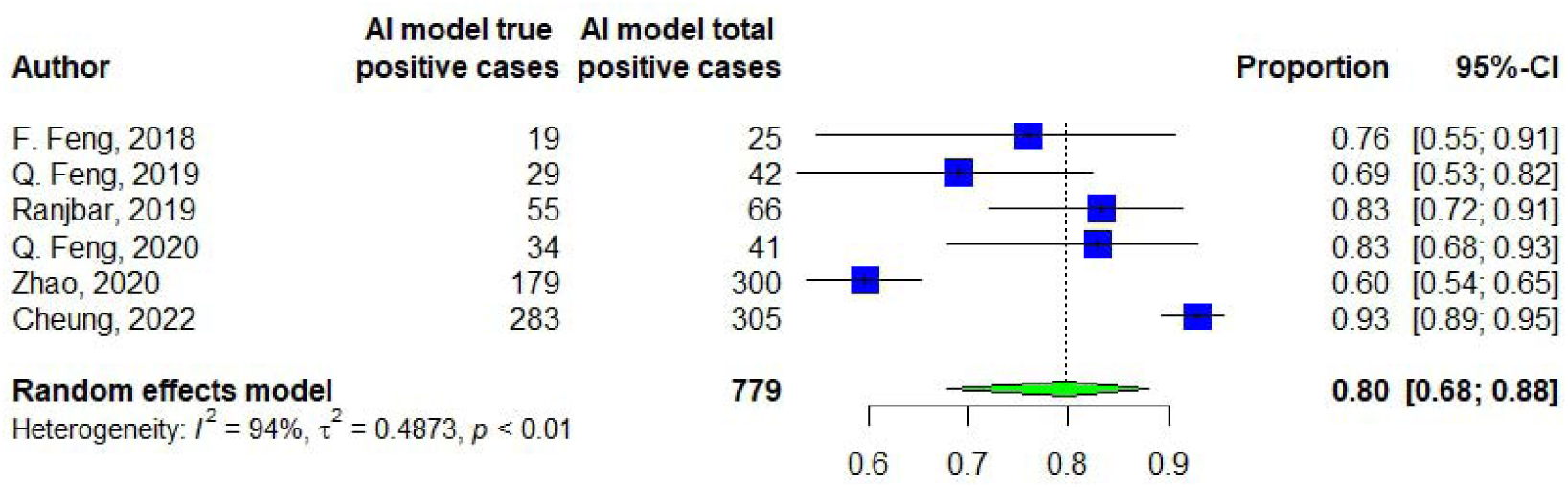
Precision of differentiating CN from MCI.

**Figure 16.**
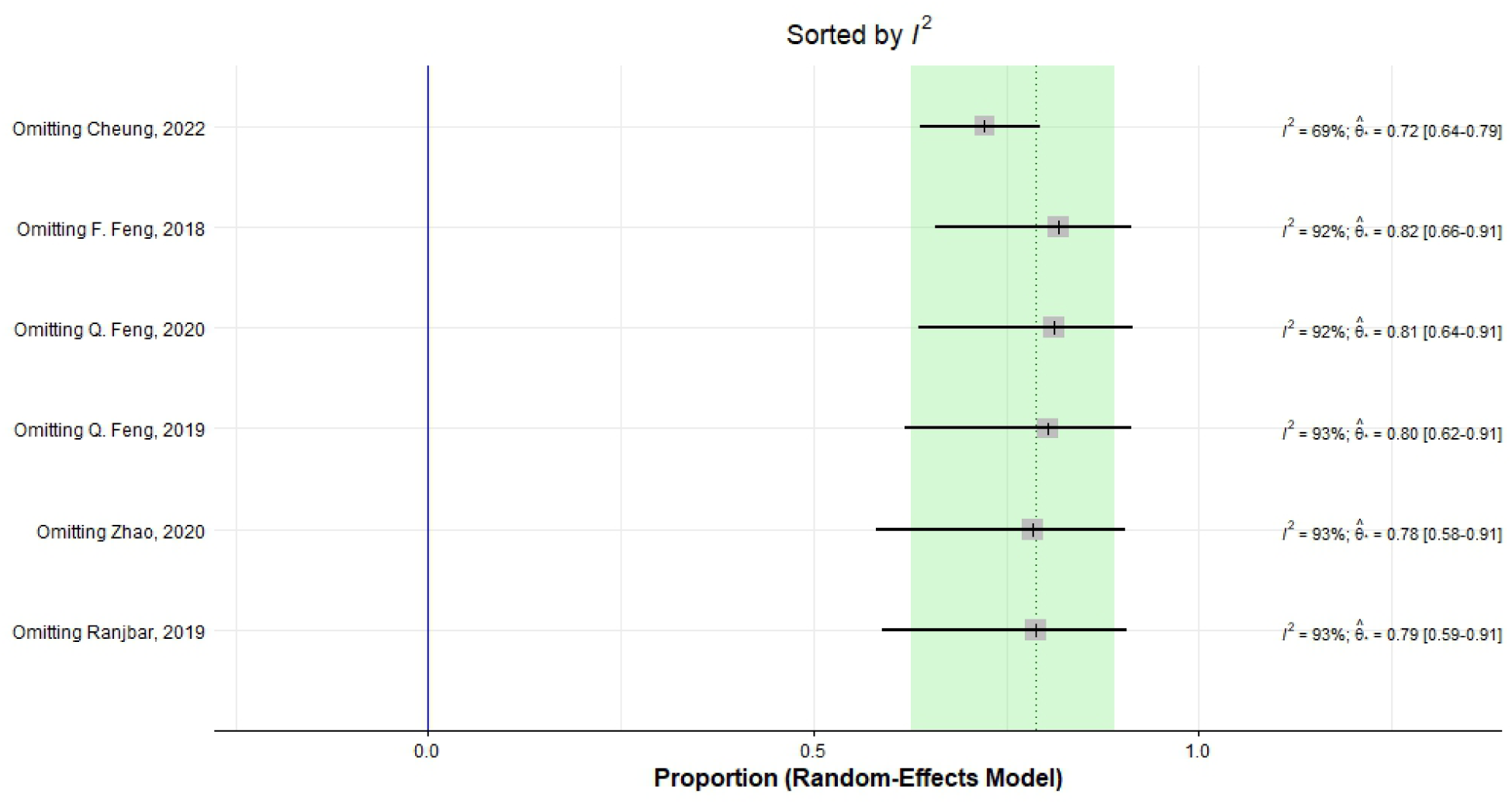
Heterogeneity of sensitivity of differentiating CN from MCI.

**Figure 17.**
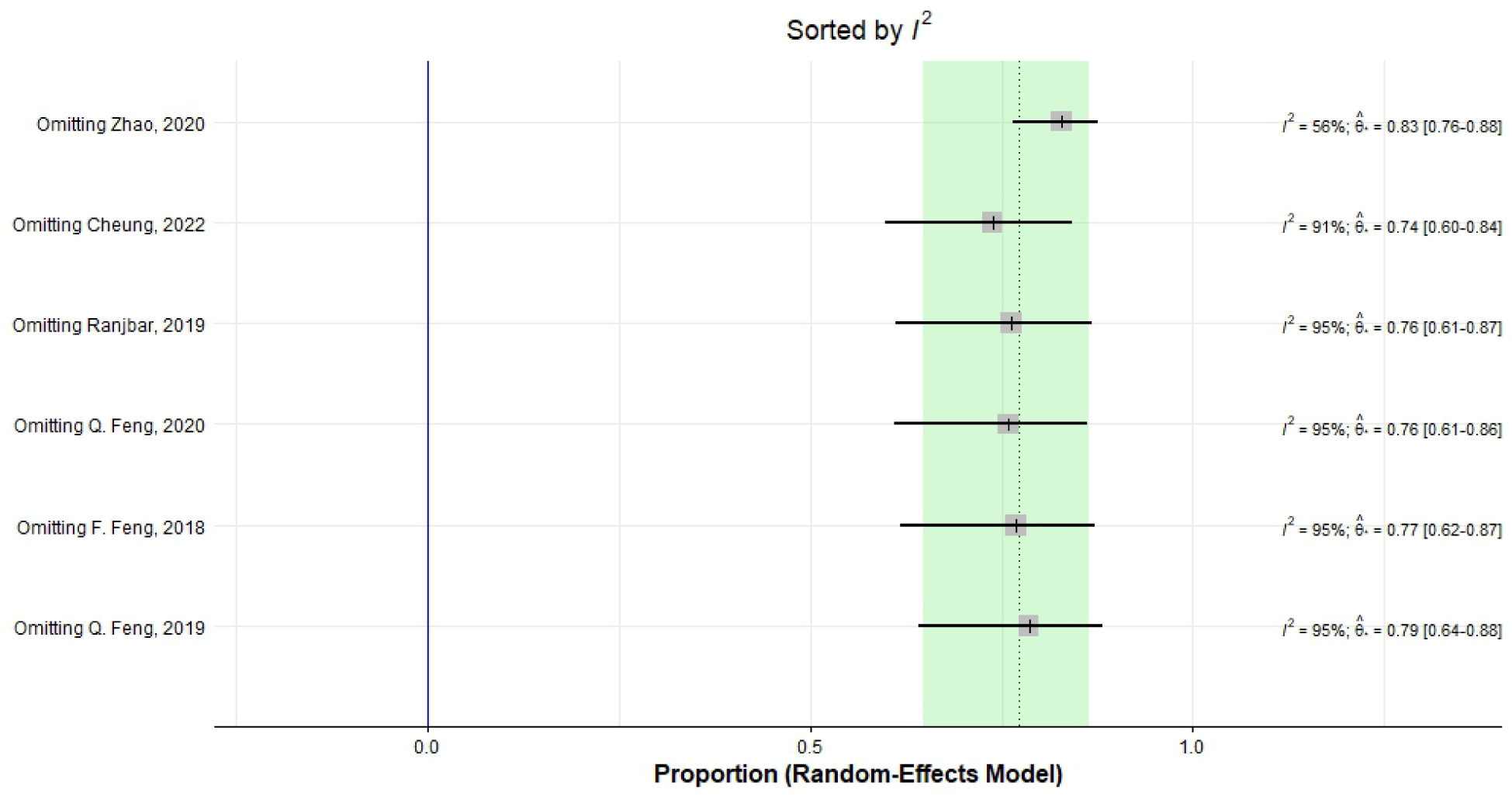
Heterogeneity of specificity of differentiating CN from MCI.

**Figure 18.**
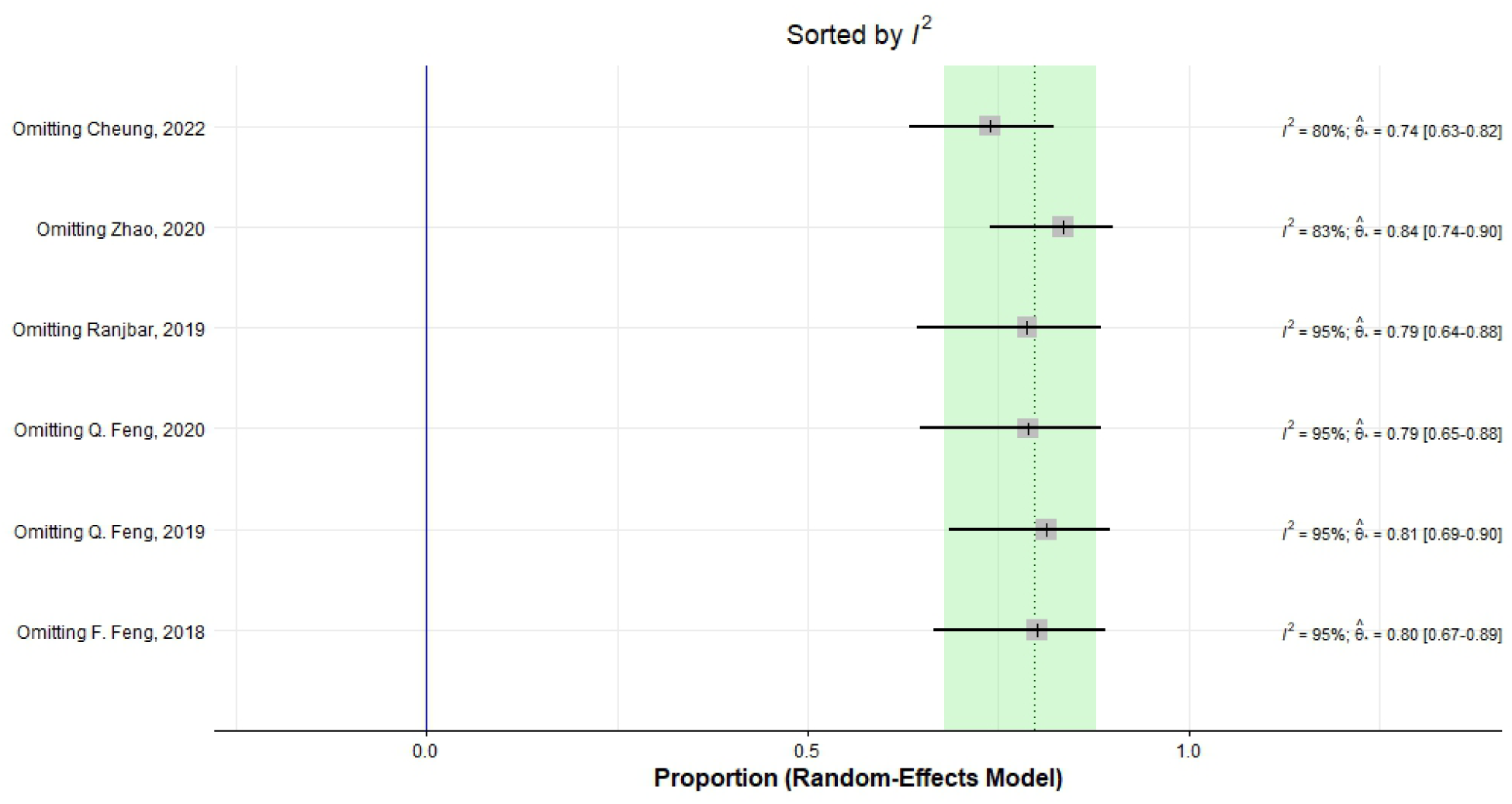
Heterogeneity of precision of differentiating CN from MCI.

## 4. Discussion

The present study, aiming to evaluate the diagnostic performance of MRI radiomics in the classification of CN, MCI, and AD, demonstrated that MRI radiomics could be considered a valuable and reliable neuroimaging biomarker for distinguishing and diagnosing these cognitive diseases, with an excellent performance of 90.4% for differentiating CN vs. AD, followed by 79.9% for CN vs. MCI, and 77.5% for MCI vs. AD. To the best of our knowledge, this is the first systematic review and meta-analysis that comprehensively evaluated the existing literature on the application of MRI radiomics and its diagnostic value in cognitive disease diagnosis. The present study suggests that radiomics can be considered a beneficial tool for the diagnosis of MCI and AD which is associated with high diagnostic accuracy and can help to improve early diagnosis, timely management, and planning of personalized treatment for individuals suffering from underlying cognitive disorders.

With a more than 2-fold increase in the number of cases since 1990, AD and other dementias affected 43.8 million people and were the fifth leading cause of mortality worldwide, with 2.4 million deaths in 2016 (Nichols et al., 2019). Besides this increasing trend in recent decades, AD incidence and burden are expected to continue to rise, partly due to increased population growth and aging (Nichols, et al., 2019). Although not all MCI patients develop AD, they are at a greater risk of progression to AD in the future. A recent systematic review and meta-analysis found that AD rate among patients suffering from MCI was 28% (Hu et al., 2017). Thus, early diagnosis and appropriate treatment of MCI can lead to a reduction in the risk of AD development in the future. Moreover, timely diagnosis and management of AD may be beneficial in lowering the disease burden.

Currently, a combination of clinical, histopathological, and radiological studies and CSF analysis are implemented to establish the MCI and AD diagnosis; however, these available methods have low sensitivity, and some cannot be routinely used in daily practice. With the purpose of improving diagnostic accuracy, prediction, and classification of diseases, radiomics were developed and became increasingly used in various clinical settings (Kumar et al., 2012).

In recent years, radiomics has shown promising results in the diagnosis of various neuropsychological disorders, such as autism spectrum disorder and attention deficit hyperactivity disorder (Chaddad, Desrosiers, & Toews, 2017; Sun et al., 2018). With an increasing trend in radiomics application in MCI and AD settings, considerable improvements in the prediction, classification, and diagnosis of these diseases have occurred. By using a quantitative approach and extracting numerous features from brain images of patients suffering from MCI and AD, radiomics can play an important role in personalized and precision medicine (Avanzo, Stancanello, & El Naqa, 2017; Lambin, et al., 2017).

By providing valuable information on microstructural changes of brain texture, radiomics gives the opportunity for early diagnosis and timely treatment of MCI at the early stages which can further help to prevent its progression towards AD. The current study found that MRI radiomics had a relatively high diagnostic accuracy in differentiating CN and MCI, with an AUC of almost 80%. Studies showed that MRI radiomics can play an important role in distinguishing individuals with CN and MCI, especially those in whom structural changes cannot be detected easily and volumetry provides limited diagnostic information (Cheung, et al., 2022). Although findings of the present study revealed that MRI radiomics had the lowest performance (77.5%) in distinguishing MCI from AD, the diagnostic value was still remarkable, with a sensitivity of almost 70%, precision, accuracy, and AUC over 74%, and specificity of almost 82%. This may represent that differentiating between MCI and AD is more difficult and challenging than distinguishing MCI from CN or AD from CN. This may be partly due to the fact that the severity of abnormal changes in brain in MCI is lower than those in AD which can make the disease diagnosis difficult.

Combining radiomics with other clinical, histopathological, laboratory, and genetic findings can help to improve the ability to distinguish MCI and AD, however, further studies are needed to assess the diagnostic value of radiomics when combined with other clinical parameters.

Our study revealed that MRI radiomics had an excellent diagnostic performance in differentiating AD from CN individuals, with a pooled AUC of 90.4%. The hippocampus was the most frequently assessed brain region by enrolled studies. It is demonstrated that the hippocampus is the major brain area mainly affected during AD (Halliday, 2017). Radiological studies revealed various degrees of hippocampal volume reduction and atrophy in MCI and AD (Catani, Dell’Acqua, De Schotten, & Reviews, 2013). In addition to structural changes, altered metabolism, abnormal function, and impaired microstructures of the hippocampus are also detected in AD (Hondius et al., 2016; Huijbers et al., 2015). Furthermore, pieces of evidence demonstrated that the changes in the hippocampus structure and texture were associated with memory performance in MCI and AD (Christensen et al., 2015; Sørensen et al., 2017; Zhang et al., 2012). Thus, the assessment of changes in the hippocampus size, structure, and function can be a useful marker for the evaluation of patients with possible underlying MCI and AD (Du et al., 2022). Although evaluating hippocampal size and volume can help diagnose AD, several recent studies showed that assessment of hippocampal texture using radiomics is more beneficial than and superior to volumetry of the hippocampus (Luk et al., 2018; Sørensen et al., 2016); however, other studies found comparable diagnostic performance between volumetry and texture analysis (Cheung, et al., 2022; Ranjbar et al., 2019).

Moreover, another beneficial advantage of radiomics is that it is useful in providing information regarding altered metabolic features in the hippocampus, such as neurofibrillary tangle and amyloid beta deposition, which affect tissue texture but cannot be detected in volumetry or MRI evaluation (Hwang et al., 2016; Manning et al., 2015). Rather than the hippocampus, other brain areas can also be affected in MCI and AD. Corpus callosum, amygdala, thalamus, and ventricles were also found to be affected in MCI and AD (Cheung, et al., 2022; De Oliveira et al., 2011; Q. Feng, et al., 2018). Moreover, a study also showed that changes may even vary in different subregions of the hippocampus and are associated with cognitive ability of individuals (F. Feng et al., 2018). Although the majority of studies focused on evaluating hippocampus texture, radiomics findings of other brain areas are studied to a less extent and need further evaluation.

The present study has several limitations that need to be addressed. First, the majority of included studies had small sample sizes, which might significantly affect the model reliability. Second, a small proportion of studies reported no external validation, which means the model generalizability cannot be ascertained. Third, in most of the studies, participant groups were not fully matched, and no adjustment for possible confounders was conducted. Fourth, although the association between radiomics findings and clinical cognitive features of patients suffering from MCI and AD has been assessed by several studies, data regarding its relationship with metabolic, genetic, and functional features is very scarce, and the possible association needs to be elaborated. Finally, since the majority of studies evaluated structural MRI radiomics and the hippocampus was mostly studied, data regarding other imaging modalities such as functional MRI, parametric MRI, and positron emission tomography, and other cerebral regions were limited and scarce. Future longitudinal, prospective, multicentric studies with larger sample sizes are needed to validate current findings, test the validity of available models, and evaluate abnormal changes in other cerebral regions during MCI and AD progression. Moreover, the applicability of using MRI radiomics in daily clinical practice needs to be understood. In addition, assessing radiomics findings of imaging modalities rather than MRI is encouraged. Lastly, the diagnostic performance of the combination of radiomics with genetic, metabolic, and laboratory markers in the screening, diagnosis, and monitoring of cognitive diseases need further investigation.

## 5. Conclusion

The present study revealed that the evaluation of brain structure and texture using MRI radiomics could be considered a useful tool for diagnosis and distinguishing MCI and AD. A growing body of literature showed that MRI radiomics, as a quantitative, non-invasive neuroimaging biomarker, had acceptable diagnostic performance in the diagnosis of MCI and AD, with high sensitivity, specificity, and accuracy. Quantitative assessment of brain texture using radiomics can lead to an improvement in MCI and AD diagnosis, planning personalized treatment, and a further reduction in the burden of these cognitive diseases. To the best of our knowledge, this is the first systematic review and meta-analysis that comprehensively reviewed and assessed the diagnostic value of MRI radiomics in MCI and AD diagnosis. Although MRI radiomics showed promising results in improving the diagnosis of these diseases, its application in daily clinical practice is still limited and needs to be proven. Moreover, it is still unknown whether combining MRI radiomics with other clinical, functional, metabolic, genetic, or laboratory markers can help to improve diagnostic performance. Moreover, the application and performance of MRI radiomics in screening and follow-up of patients with underlying MCI and AD are yet to be understood.

## Supporting information

Keywords

Search Queries

## Data Availability

All data produced in the present study are available upon reasonable request to the authors

## Acknowledgment

The authors appreciate Mohammad Amin Habibi for his kind help during the manuscript preparation

---

**Supplementary Figure 1.**
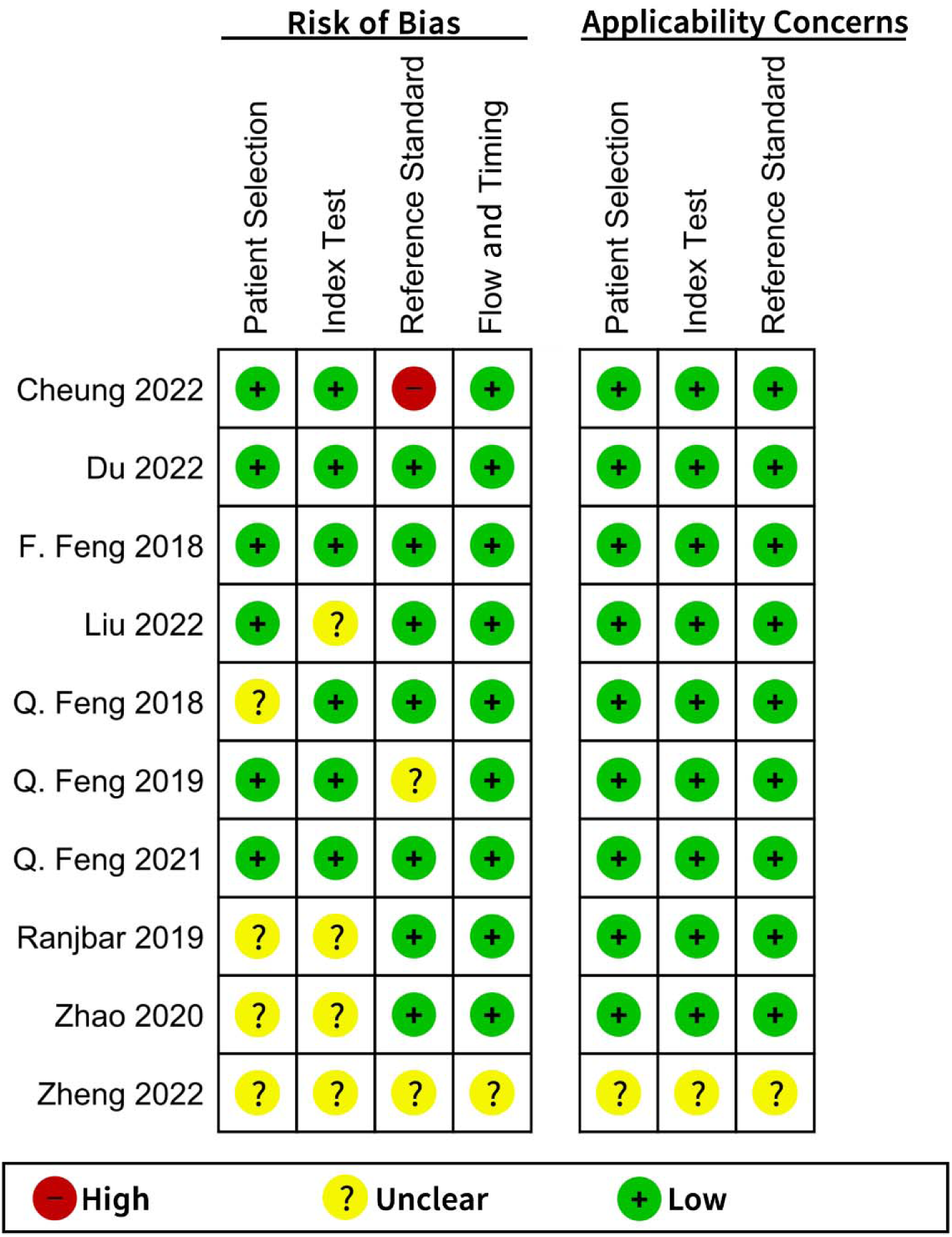
Critical appraisal based on the QUADAS tool - part 1.

**Supplementary Figure 2.**
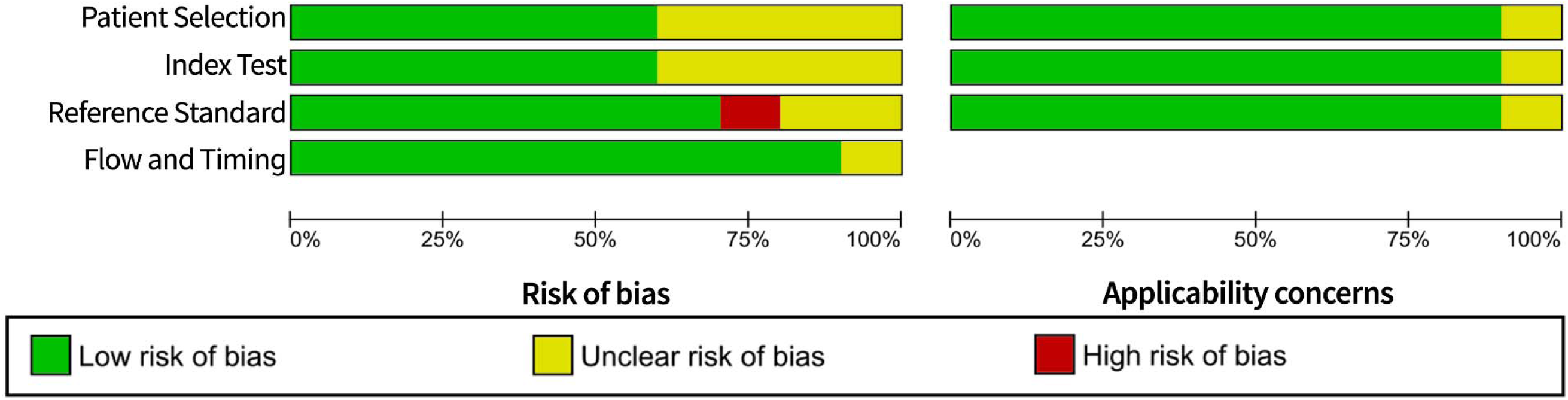
Critical appraisal based on the QUADAS tool - part 2.

**Supplementary Figure 3.**
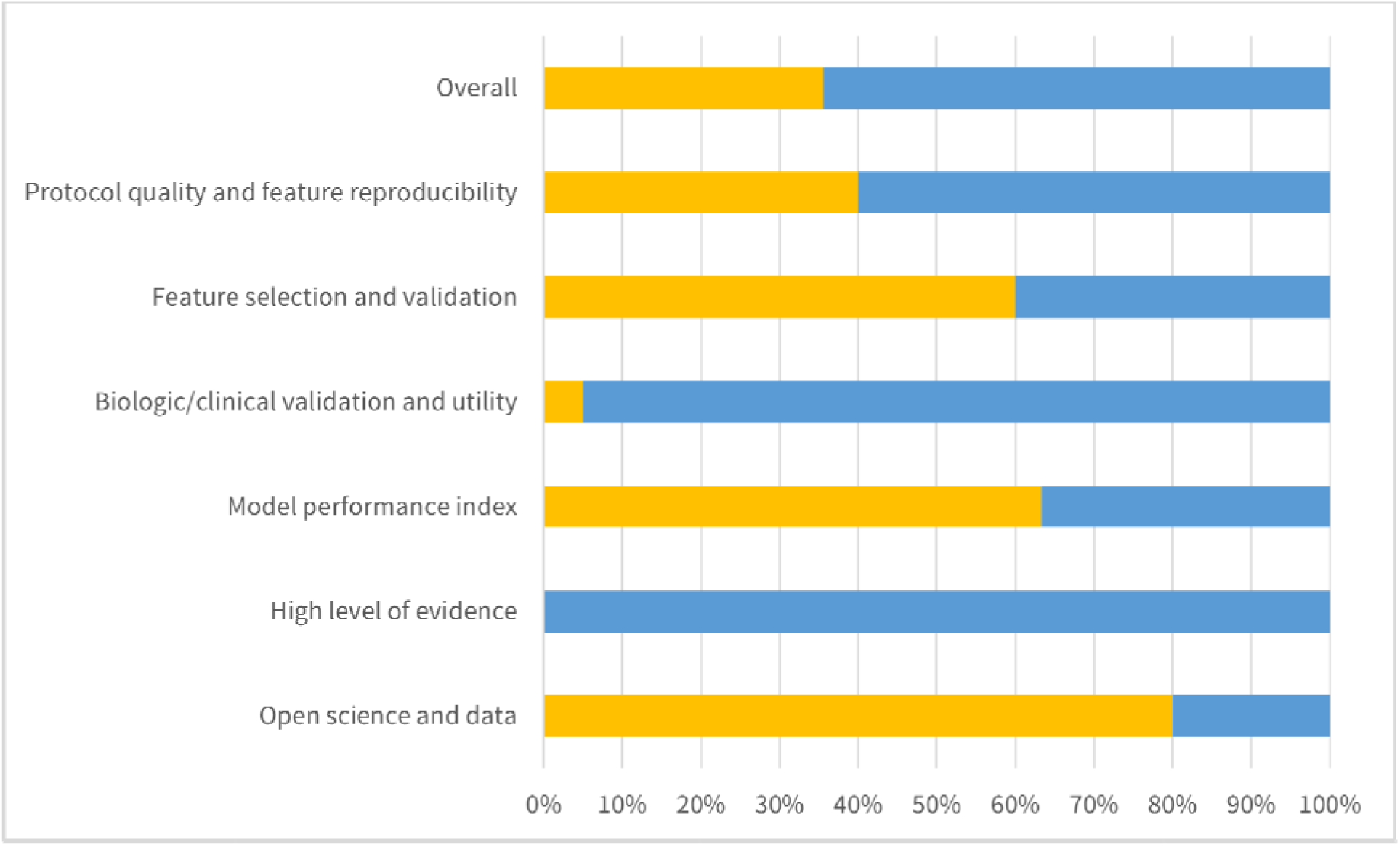
Critical appraisal based on the RQS tool.

**Supplementary Table 1.** Basic adherence rate according to the six key domains.

|  | Basic adherence rate |
| --- | --- |
| Total 16 items | 35.6% |
| Domain 1: Protocol quality and stability in image and segmentation | 40% |
| Protocol quality | 10 (100%) |
| Test-retest | 5 (50%) |
| Phantom study | 1 (10%) |
| Multiple segmentation | 0 (0%) |
| Domain 2: Feature selection and validation | 60% |
| Feature reduction or adjustment of multiple testing | 7 (70%) |
| Validation | 5 (50%) |
| Domain 3: Biologic/clinical validation and utility | 5% |
| Multivariate analysis with non-radiomics features | 0 (0%) |
| Biologic correlates | 1 (10%) |
| Comparison to 'gold standard' | 1 (10%) |
| Potential clinical utility | 0 (0%) |
| Domain 4: Model performance index | 63.3% |
| Discrimination statistics | 10 (100%) |
| Calibration statistics | 9 (90%) |
| Cut-off analysis | 0 (0%) |
| Domain 5: High level of evidence | 0% |
| Prospective study | 0 (0%) |
| Cost-effective analysis | 0 (0%) |
| Domain 6: Open science and data | 8 (80%) |

**Supplementary Table 2.**
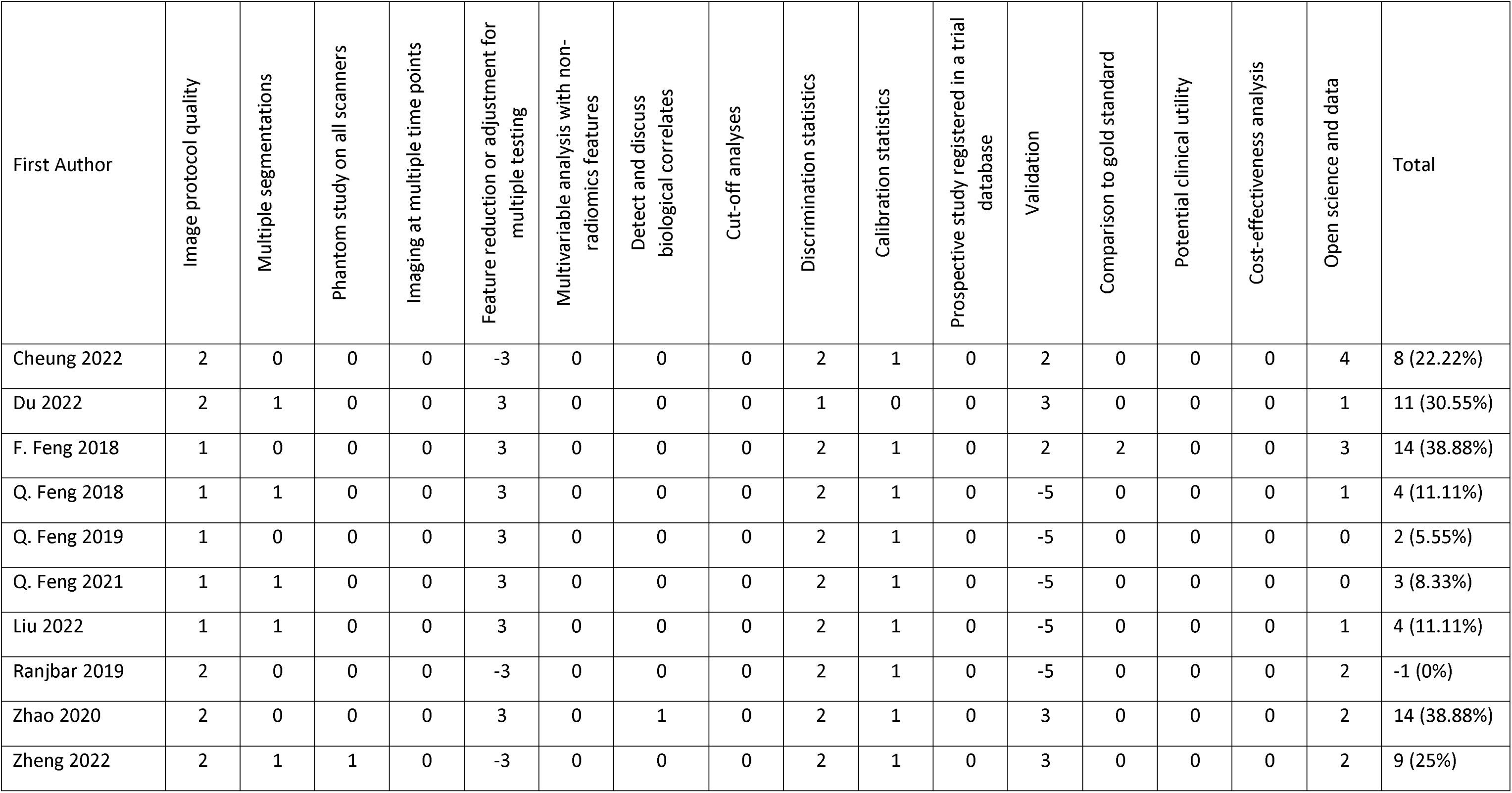
Details of quality assessment by Radiomics Quality Score (RQS) of all included studies.

