## Supplementary material for "Diagnostic Performance of MRI Radiomics for Classification of Alzheimer’s disease, Mild Cognitive Impairment, and Normal Subjects: A Systematic Review and Meta-analysis": Keywords

“Alzheimer Keywords”

MESH Terms:

Alzheimer Disease

Emtree Terms:

Alzheimer disease

SYNONYMS:

Alzeimer

Alzheimer

Primary Senile Degenerative Dementia

Senile Dementia

Presenile Dementia

Mild Cognitive Impairment

“Radiomic Keywords”

MESH Terms:

.

Emtree Terms:

Radiomics

SYNONYMS:

Radiomics

Radiomic
