## Supplementary material for "Diagnostic Performance of MRI Radiomics for Classification of Alzheimer’s disease, Mild Cognitive Impairment, and Normal Subjects: A Systematic Review and Meta-analysis": Search Queries

Pubmed Search Query

Time of search: 17 October 2022
Results: 46

(“Alzheimer Disease”[mesh] OR Alzeimer[tiab] OR Alzheimer[tiab] OR Primary Senile Degenerative Dementia[tiab] OR Senile Dementia[tiab] OR Presenile Dementia[tiab] OR Mild Cognitive Impairment[tiab]) AND (Radiomics[tiab] OR Radiomic[tiab])

Embase Search Query

Time of search: 17 October 2022
Results: 65

(‘Alzheimer disease’/exp OR ‘Alzeimer’:ab,ti OR ‘Alzheimer’:ab,ti OR ‘Primary Senile Degenerative Dementia’:ab,ti OR ‘Senile Dementia’:ab,ti OR ‘Presenile Dementia’:ab,ti OR ‘Mild Cognitive Impairment’:ab,ti) AND (‘Radiomics’/exp OR ‘Radiomics’:ab,ti OR ‘Radiomic’:ab,ti)

Scopus Search Query

Time of search: 17 October 2022
Results: 62

(TITLE-ABS (“Alzeimer” OR “Alzheimer” OR “Primary Senile Degenerative Dementia” OR “Senile Dementia” OR “Presenile Dementia” OR “Mild Cognitive Impairment”)) AND (TITLE-ABS (“Radiomics” OR “Radiomic”))

Web of science Search Query

Time of search: 17 October 2022
Results: 61

(TS= (“Alzeimer” OR “Alzheimer” OR “Primary Senile Degenerative Dementia” OR “Senile Dementia” OR “Presenile Dementia” OR “Mild Cognitive Impairment”)) AND (TS= (“Radiomics” OR “Radiomic”))
